# Genomic insights into methicillin-resistant *Staphylococcus aureus* from Swiss wastewater and clinical samples

**DOI:** 10.64898/2026.09.17.26361412

**Authors:** Julia Kamer, Sheena Conforti, Silvio D. Brugger, Patrice François, Valeria Gaia, Tanja Stadler, Stephan Harbarth, Timothy R. Julian

## Abstract

**Objectives:** To determine whether methicillin-resistant *Staphylococcus aureus* (MRSA) recovered from wastewater, as a proxy for community carriage, and clinical settings represent distinct populations and whether their comparison provides insight into distinctions between healthcare-associated (HA-) and community-associated (CA-) MRSA.

**Methods:** We analyzed whole-genome sequences from 115 MRSA isolates collected in Switzerland between 2021 and 2024, including 89 from three hospitals and 26 from six wastewater treatment plants serving the hospital catchment areas. We compared the phylogenetic relatedness, sequence types (STs), SCC*mec* types, *spa* types, antimicrobial resistance genes (ARGs), and virulence factors between the two sources.

**Results:** Amongst all isolates, few were closely genetically related, with only 5% (273/6,105) of the pairwise comparisons differing by ≤15 core-genome single nucleotide polymorphisms. Amongst the 89 clinical isolates, there were 23 STs and 46 *spa* types as compared to 6 STs and 9 *spa* types amongst the 26 wastewater isolates. All wastewater isolates (100%, or 26/26) and most clinical isolates (64%, or 57/89) were SCC*mec* IV, historically associated with CA-MRSA. Panton-Valentine Leukocidin genes, which historically indicated virulence associated with CA-MRSA, were detected in 37% (43/115) of isolates, including 40% (36/89) of clinical isolates and 27% (7/26) of wastewater isolates. One wastewater ST (ST97, 6/26 isolates) was phylogenetically distinct from all clinical isolates and corresponds to a known livestock-associated MRSA lineage, while other STs identified in wastewater are known to circulate in livestock or communities (ST22, ST88, ST1482).

**Conclusions:** Our findings highlight the effectiveness of clinical surveillance in monitoring MRSA epidemiology in Switzerland, with most major lineages, antimicrobial resistance determinants, and virulence factors identified in wastewater also present among clinical isolates. Notably, wastewater isolates shared sequence types with known livestock and CA-MRSA lineages. This suggests wastewater supplements traditional methods in understanding community circulation but provides limited unique insight into clinical circulation.

## Introduction

*Staphylococcus aureus* is a major cause of community- and healthcare-associated infections (1,2). Methicillin-resistant *S. aureus* (MRSA) is a high-priority pathogen that causes more than 100,000 deaths annually worldwide (3,4). Resistance to methicillin is mediated by the *mecA* or *mecC* gene carried on the staphylococcal cassette chromosome *mec* (SCC*mec*), a mobile genetic element that contributes to the spread of resistance among staphylococci (5).

Historically, MRSA epidemiology has been divided into healthcare-associated (HA-MRSA), community-associated (CA-MRSA), and livestock-associated (LA-MRSA). HA-MRSA and CA-MRSA are distinguished by different SCC*mec* types, virulence factors, and antimicrobial resistance profiles (6). HA-MRSA has been associated with SCC*mec* types I-III and broader antimicrobial resistance profiles, whereas CA-MRSA with SCC*mec* types IV-V and virulence factors such as Panton-Valentine Leukocidin (PVL) (6). Increasing detection of SCC*mec* IV MRSA in hospitals and rising multidrug resistance among CA-MRSA lineages have blurred the traditional HA/CA distinction (7,8). MRSA surveillance is largely based on isolates recovered from healthcare settings. Consequently, the diversity of MRSA circulating outside hospitals may be underrepresented, limiting our understanding of how CA-/HA-MRSA populations overlap.

Wastewater-based surveillance has emerged as a complementary approach to clinical surveillance because it provides a composite representation of bacterial populations circulating within large urban catchments (9,10). Recent studies have recovered MRSA and other staphylococci from wastewater and used whole-genome sequencing (WGS) to characterize their clonal diversity, antibiotic resistance determinants, and mobile genetic elements (11,12). Conceptually, urban wastewater is a potential source of CA-MRSA from people who do not visit clinics but shed it into the wastewater, as well as HA-MRSA from patients living in the catchment and from discharged hospital wastewater. As such, wastewater may capture MRSA lineages circulating outside healthcare settings, whereas clinical isolates are more likely to reflect healthcare-associated lineages. However, wastewater may also capture pathogens circulating in livestock and wildlife. It is therefore unclear to what extent wastewater and clinical surveillance capture overlapping MRSA populations, and whether wastewater-derived MRSA can provide additional insights that support clinical MRSA epidemiology, including the distinction between CA- and HA-MRSA.

In this study, we compared whole-genome sequences of 115 MRSA isolated from wastewater and geographically linked hospitals in Switzerland from 2021 to 2024. Specifically, we *i)* assessed the genetic relatedness of wastewater and clinical MRSA populations using phylogenetic analyses, *ii)* compared their genomic characteristics, including sequence types (STs), SCC*mec* types, and *spa* types, and *iii)* compared molecular signatures associated with antimicrobial resistance and virulence across sampling sources.

## Methods

### Collection of clinical isolates

Between April 2021 and June 2024, 90 MRSA isolates were obtained from three Swiss healthcare facilities: Ente Ospedaliero Cantonale (EOC, Lugano; n=31), University Hospital of Geneva (HUG, Geneva; n=29), and University Hospital Zurich (USZ, Zurich; n=30) (**Table S1, Figure S1**). Isolates included invasive MRSA from hospitalized patients and routine screening specimens and were selected to represent the diversity of MRSA circulating at each institution during the study period, as previously described (13). Samples were transported to the laboratory in eSwab tubes (Copan, USA) or on blood agar plates, stored at 4°C on arrival, and processed the next day by streaking onto LB agar (Lennox) and incubating at 37°C for 24h. Single colonies were inoculated into Luria Broth (AppliChem) and incubated at 37°C overnight before DNA extraction.

### Collection of wastewater isolates

Composite influent wastewater was collected every four weeks from December 2022 to January 2024 at six wastewater treatment plants (WWTPs) in Switzerland (**Figure S1**) (14). Presumptive MRSA were isolated by plating 100µL of undiluted wastewater onto CHROMagar MRSA chromogenic media (CHROMagar, France), followed by incubation at 37°C for 24h. At each time point and WWTP, up to seven pink-to-mauve colonies were randomly selected, streaked onto LB agar (Lennox), and incubated at 37°C for 24h. Single colonies were then inoculated into 200µL Luria Broth (AppliChem) and incubated at 37°C for 24h. Glycerol stocks were prepared at a final concentration of 12% and stored at −80°C.

### PCR confirmation of presumptive MRSA wastewater isolates

To confirm the identity of presumptive MRSA isolates recovered from wastewater, DNA was extracted by microwave lysis by resuspending a single colony in 20 µL of nuclease-free water, vortexing, and heating at 900 W for 5 minutes (15). DNA was screened for *S. aureus* using the 16SSA PCR targeting a single-base-pair mismatch in the 16S ribosomal RNA gene (16).

### DNA extraction and whole genome sequencing

To obtain high-quality genomic DNA for whole-genome sequencing (WGS), DNA was extracted from 0.9 mL overnight cultures of both wastewater and clinical MRSA isolates using the DNeasy Blood & Tissue kit (Qiagen, USA) following the manufacturer’s instructions and eluted in 100µL Buffer AE. DNA concentration and purity were measured by spectrophotometry (NanoDrop; Thermo Fisher Scientific). DNA extracts were shipped on ice to the Earlham Institute (Norwich, UK), where libraries were prepared using the LITE pipeline and sequenced on a NovaSeqX 1.5B flow cell to generate 150 bp paired-end reads (17).

### Bioinformatic analyses and isolate characterization

All bioinformatic analyses were implemented using a Snakemake workflow and are available at https://github.com/sheenaconforti/ecoli-wgs. Illumina reads were quality-filtered, *de novo* assembled, and annotated (**Supplementary Material 1**). A recombination-corrected core-genome alignment was used to reconstruct a maximum-likelihood phylogeny and calculate pairwise core-genome single-nucleotide polymorphism (cgSNP) distances. Clonal isolate pairs with 0 cgSNPs difference recovered from the same wastewater sampling location and time point were collapsed to single representative isolates (**Table S2**). Recent transmission was inferred using a threshold of ≤15 cgSNPs (18). Isolates were characterized by sequence type (ST), SCC*mec* type, *spa* type, antimicrobial resistance genes (ARGs), and virulence factors (VFs) (**Supplementary Material 2**). ARGs were assigned to 14 clinically relevant antimicrobial classes (**Table S3**). *Spa* typing was additionally performed by PCR amplification and Sanger sequencing of the *spa* gene (**Supplementary Material 3**). Statistical differences in the number of ARGs and antimicrobial resistance classes detected per isolate between clinical and wastewater sources were assessed using the Mann-Whitney U test with α < 0.05. To compare the overall diversity of ARGs between sources while accounting for unequal sample sizes, rarefaction was performed by randomly selecting 26 distinct clinical isolates across 10,000 iterations and counting the number of unique ARGs detected among the selected isolates. Statistical analyses were performed in R (v4.4.1) and RStudio (v2024.04.2).

## Results

### Isolates recovery and WGS

Of 512 presumptive MRSA isolates recovered from wastewater, 27 (5%) were confirmed as *S. aureus* by the 16SSA PCR assay (**Table S4**). WGS was performed on 117 MRSA isolates, comprising 27 wastewater and 90 clinical isolates. After excluding one low-coverage sample and one sample lacking *S. aureus* reads, 115 genomes (26 wastewater and 89 clinical isolates) with coverage ranging from 192× to 844× were retained for downstream analyses (**Table S5**).

### Genetic relatedness between clinical and wastewater MRSA isolates

The lineages captured in wastewater only partially overlapped with those detected among clinical isolates. We observed more lineages in clinical isolates (23 STs) than in wastewater (6 STs), as expected from differences in sample sizes. Amongst the lineages found in wastewater, all except one (5 of 6 STs) were also found in the clinical isolates (**Figure 1**).

**Figure 1:**
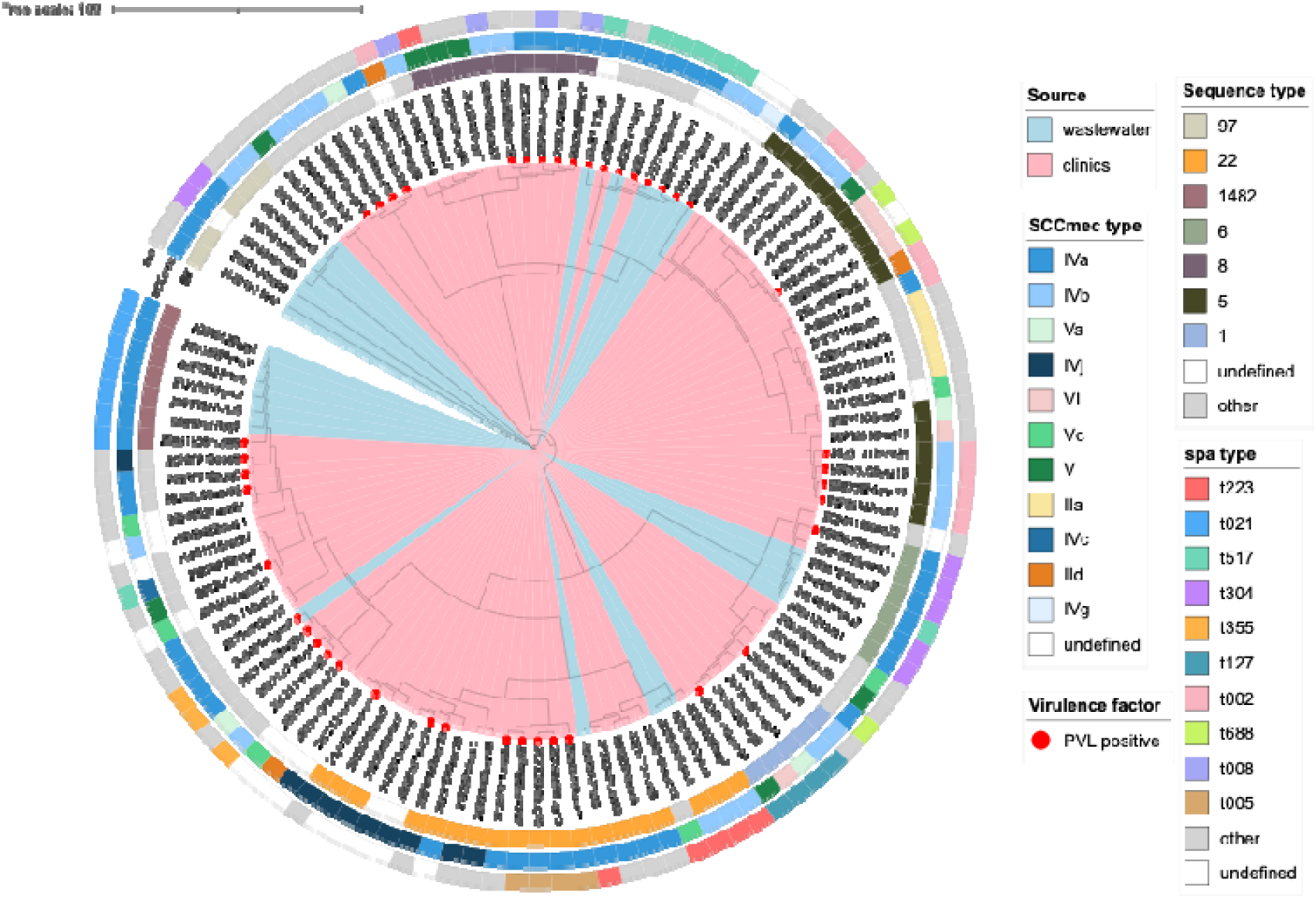
Phylogenetic tree of 115 methicillin-resistant *S. aureus* (MRSA) isolates from wastewater (n=26) and clinical cultures (n=89), inferred from the recombination-masked coregene alignment. Tip labels of wastewater isolates encode the isolation date, site and a within-date/location index (to distinguish isolates from the same time point and location), labels of clinical isolates encode the isolation date, the hospital, and a within-location only index. Annotations illustrate the distribution of sequence type (ST), SCC*mec* types, spa types, and sources of collection (wastewater or clinics). STs are colored if present in five or more isolates, and spa types are colored if present in three or more isolates. Other, less frequent STs and spa types are colored in grey, and undefined types in white. Isolates positive for Panton-Valentine Leukocidin (PVL), defined as carriage of both the lukF-PV and lukS-PV genes at ≥80% sequence identity, are marked with a red dot between the outer ring and the tip label.

Using a threshold of ≤ 15 cgSNPs to indicate transmission within the previous six months, 273 genetically related isolate pairs were identified among 6,105 possible pairwise comparisons (5%) (**Table 1**). This corresponded to 197/3,916 (5%) comparisons between clinical isolates, 26/231 (11%) comparisons between wastewater isolates, and 50/1,958 (3%) comparisons between clinical and wastewater isolates. Among the cross-compartment pairs, a clinical isolate from EOC Ticino was collected 12 days before its wastewater counterpart in Chur, while in Geneva the clinical isolate preceded the related wastewater isolate by 15 days.

**Table 1:**
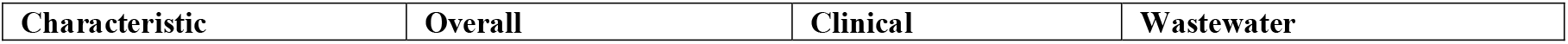
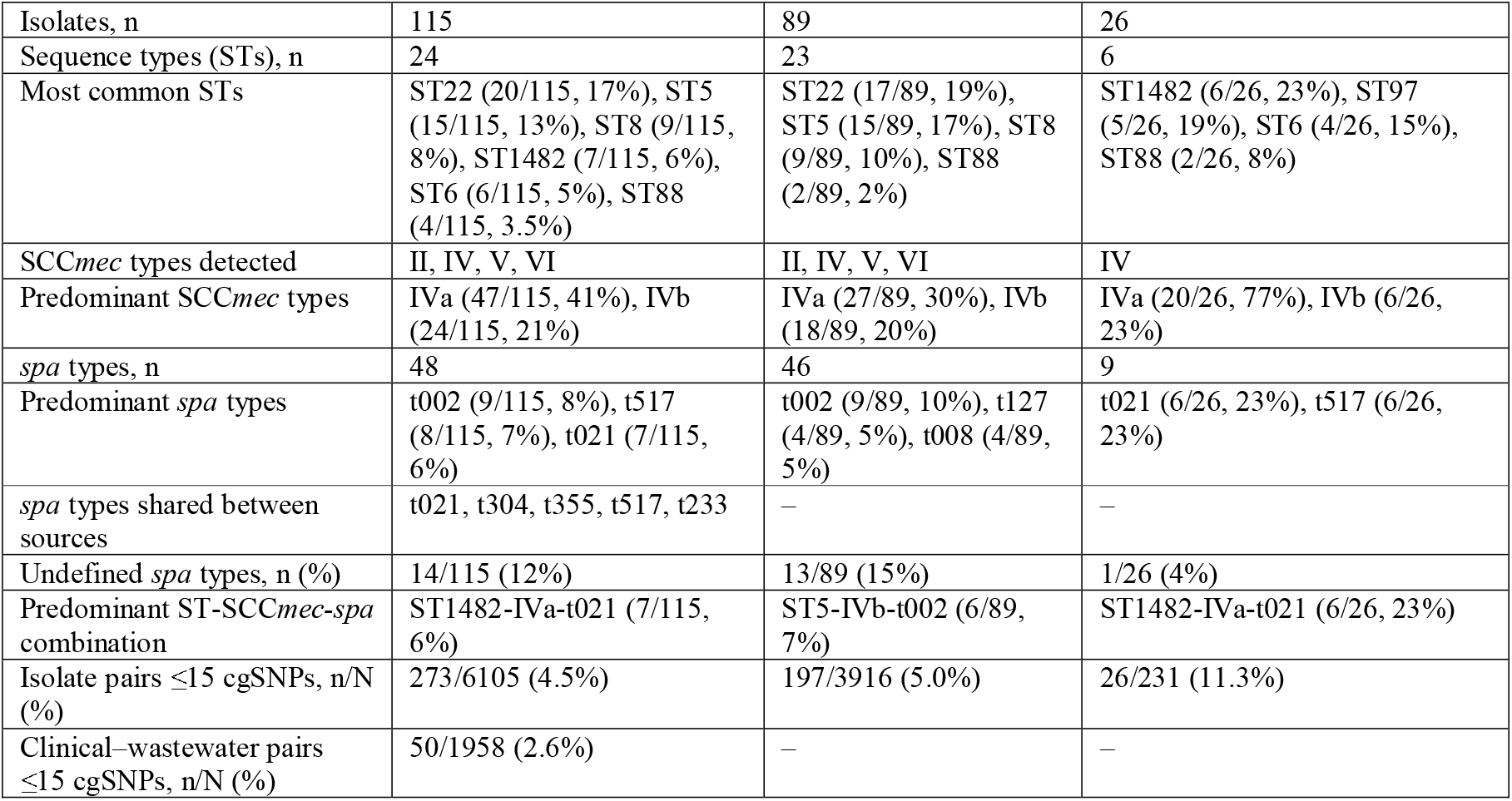
Genomic characteristics of MRSA isolates recovered from clinical and wastewater sources. Sequence type (ST), staphylococcal cassette chromosome *mec* (SCC*mec*) type, *spa* type, and pairwise core-genome single nucleotide polymorphism (cgSNP) analyses are summarized overall and by source. Values are presented as n (%) unless otherwise indicated, where n denotes the number of isolates and percentages are calculated using the total number of isolates within each source category as the denominator. For pairwise cgSNP comparisons, n/N (%) indicates the number of isolate pairs at or below a threshold of 15 cgSNPs (n) divided by the total number of pairwise comparisons (N) within each category. Shared *spa* types represent *spa* types detected in both clinical and wastewater isolates.

### Genetic diversity of MRSA

More STs were observed among clinical than wastewater isolates (**Table 1, Table S6**). In total, 24 STs were identified, with the most common being ST22, ST5, ST8, ST1482, and ST6. Lineage composition differed between sampling sources, with ST22 and ST5 predominating among clinical isolates, ST1482, ST97, and ST6 among wastewater isolates, and ST88 intermixed between clinical and wastewater (Table S6). ST97 was detected exclusively in wastewater isolates.

SCC*mec* IV predominated in both sampling sources and was the only cassette type identified among wastewater isolates (**Table 1, Table S7**). SCC*mec* types II, V, and VI were detected exclusively among clinical isolates.

Clinical isolates displayed greater *spa*-type diversity than wastewater isolates, comprising 46 and 9 spa types, respectively (**Table 1, Table S8**). Overall, 48 *spa* types were identified, but only five (t021, t304, t355, t517, and t233) were shared between clinical and wastewater isolates. Wastewater isolates were dominated by *spa* types t021 and t517, whereas the most frequent *spa* types in clinical isolates were t002, t127, and t008. No *spa* type could be assigned to 14 isolates, whether using WGS or Sanger sequencing.

Three ST-SCC*mec*-spa combinations predominated (**Table 1, Table S9**). The most frequent was ST1482-IVa-t021, detected in six wastewater isolates and one clinical isolate, followed by ST5-IVb-t002, detected exclusively among six clinical isolates, and ST6-IVa-t304, detected in two clinical isolates and three wastewater isolates. All six wastewater-associated ST1482-IVa-t021 were collected in June 2023, from three distinct WWTPs (Lugano, Chur, and Geneva), whereas the corresponding clinical isolate was recovered from a patient at HUG in December 2023.

### Antimicrobial resistance genes and virulence factors

Antimicrobial resistance profiles were similar between clinical and wastewater MRSA isolates. The prevalence of multidrug resistance (MDR; resistance to at least three antimicrobial classes) and the distribution of resistance classes were comparable between sources (**Table 2, Table S10**). No significant differences were observed between clinical and wastewater isolates in either the number of ARGs detected per isolate or the number of antimicrobial classes represented (**Figure S2**). Rarefaction analysis accounting for differences in sample size yielded similar estimates of the number of unique ARGs detected in clinical and wastewater isolates (**Table 2, Figure S3**).

**Table 2:** Genotypic antimicrobial resistance and virulence characteristics of MRSA isolates recovered from clinical and wastewater sources. Values are presented as n (%) unless otherwise indicated. Antimicrobial resistance classes were assigned based on antimicrobial resistance genes (ARGs) identified using the Comprehensive Antibiotic Resistance Database (CARD). Multidrug resistance (MDR) was defined as the presence of ARGs associated with resistance to ≥3 antimicrobial classes; efflux pumps were not considered a resistance class. Numbers of ARGs and resistance classes per isolate are reported as median (interquartile range, IQR). To account for differences in sample size between sources, clinical isolates were randomly subsampled to match the number of wastewater isolates (n = 26) across 10,000 rarefaction iterations; values represent the median number of unique ARGs detected with empirical 95% confidence intervals. Virulence factors assessed included Panton–Valentine leukocidin (PVL), enterotoxins, exfoliative toxins, and the *spa* gene.

| Characteristic | Overall | Clinical | Wastewater |
| --- | --- | --- | --- |
| Number of isolates tested, n | 115 | 89 | 26 |
| Unique ARGs detected, n | 37 | 34 | 26 |
| Rarefied unique ARGs detected, n* | - | 29 (95% CI 25-32) | 26 |
| ARGs per isolate, median (IQR) | 12 (11-13) | 12 (11-14) | 12 (11-13) |
| Antimicrobial resistance classes per isolate, median (IQR) | 5 (3-8) | 5 (3-7) | 5 (3-9) |
| Multidrug-resistant isolates, n (%) | 103 (89.6%) | 81 (91.0%) | 22 (84.6%) |
| Isolates carrying efflux pump genes, n (%) | 114 (99.1%) | 88 (98.9%) | 26 (100%) |
| β-lactam resistance genes, n (%) | 111 (96.5%) | 86 (96.6%) | 25 (96.2%) |
| Tetracycline resistance genes, n (%) | 110 (95.7%) | 84 (94.4%) | 26 (100%) |
| Fosfomycin resistance genes, n (%) | 65 (56.5%) | 49 (55.1%) | 16 (61.5%) |
| Lincosamide resistance genes, n (%) | 59 (51.3%) | 45 (50.6%) | 14 (53.8%) |
| Macrolide resistance genes, n (%) | 56 (48.7%) | 43 (48.3%) | 13 (50.0%) |
| Streptogramin resistance genes, n (%) | 56 (48.7%) | 43 (48.3%) | 13 (50.0%) |
| Diaminopyrimidine resistance genes, n (%) | 42 (36.5%) | 29 (32.6%) | 13 (50.0%) |
| Aminoglycoside resistance genes, n (%) | 40 (34.8%) | 34 (38.2%) | 6 (23.1%) |
| Fusidic acid resistance genes, n (%) | 36 (31.3%) | 30 (33.7%) | 6 (23.1%) |
| Oxazolidinone resistance genes, n (%) | 23 (20.0%) | 16 (18.0%) | 7 (26.9%) |
| Phenicol resistance genes, n (%) | 23 (20.0%) | 16 (18.0%) | 7 (26.9%) |
| Pleuromutilin resistance genes, n (%) | 23 (20.0%) | 16 (18.0%) | 7 (26.9%) |
| Nucleoside resistance genes, n (%) | 9 (7.8%) | 8 (9.0%) | 1 (3.8%) |
| Mupirocin resistance genes, n (%) | 1 (0.9%) | 1 (1.1%) | 0 (0%) |
| PVL-positive isolates, n (%) | 43 (37.4%) | 36 (40.4%) | 7 (26.9%) |
| Enterotoxin-positive isolates, n (%) | 73 (63.5%) | 61 (68.5%) | 12 (46.2%) |
| Exfoliative toxin-positive isolates, n (%) | 6 (5.2%) | 6 (6.7%) | 0 (0%) |
| spa-positive isolates, n (%) | 13 (11.3%) | 10 (11.2%) | 3 (11.5%) |

In total, 37 ARGs conferring resistance to 14 antimicrobial classes were identified across the 115 MRSA genomes (**Figure 2**). The most prevalent genes were *mepR, mgrA, arlR, mecA*, and *mepA* (**Table S11**). Genes associated with resistance to β-lactams were nearly ubiquitous, and determinants associated with tetracyclines, fosfomycin, lincosamides, macrolides, and diaminopyrimidines were also common (**Table 2**). Mupirocin resistance genes were detected in only a single clinical isolate.

**Figure 2:**
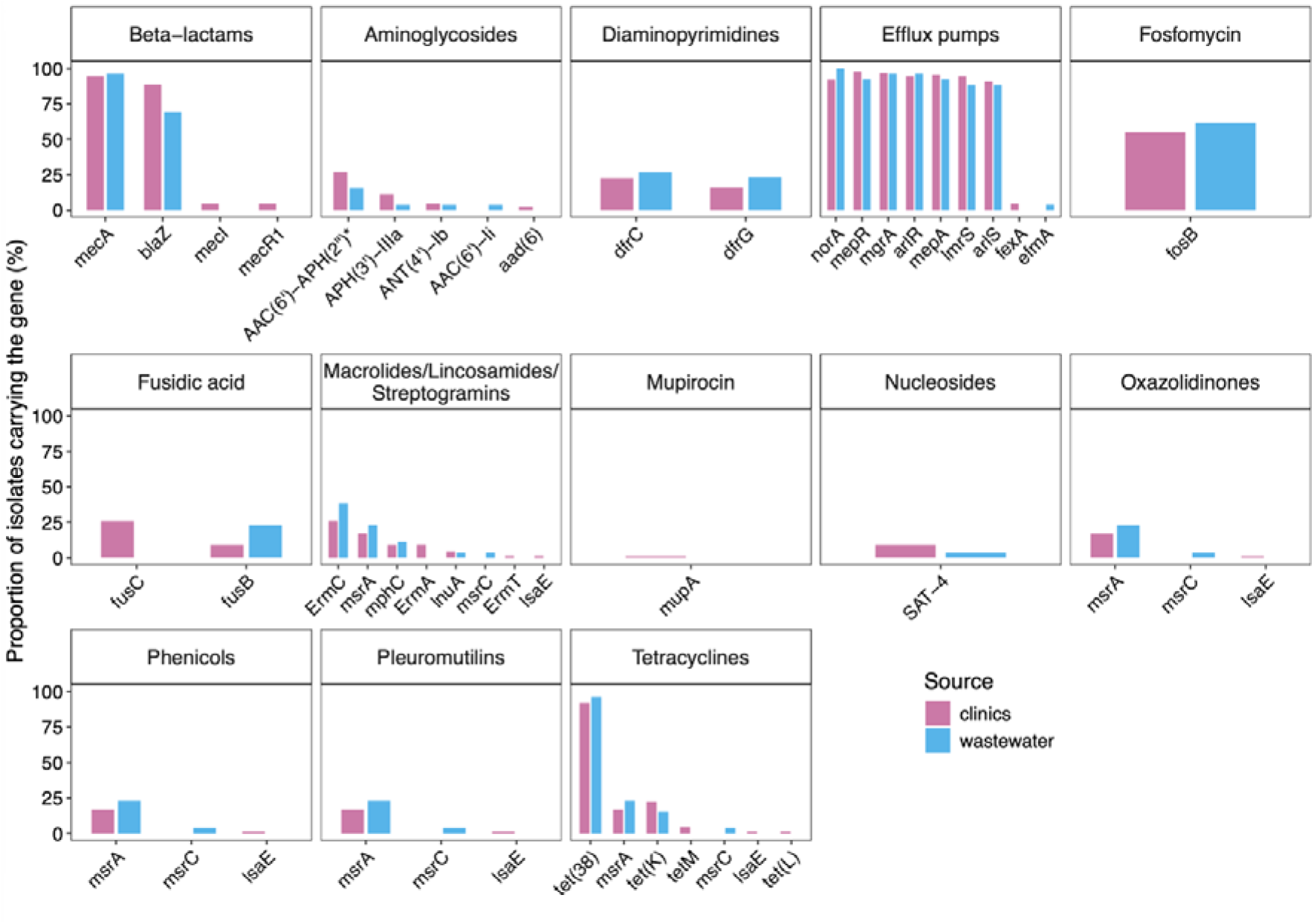
Distribution of antibiotic resistance genes across sources in MRSA isolates. The proportion of MRSA isolates carrying genes associated with different antibiotic resistance classes, based on annotations from the Comprehensive Antibiotic Resistance Database (CARD). Source is represented in colors: clinics (pink) and wastewater (blue). *AAC(6’)-APH(2’’) refers to the bifunctional aminogly-coside-modifying enzyme AAC(6’)-Ie-APH(2’’)-Ia. Each bar shows the proportion of isolates from a specific source carrying the corresponding gene.

Virulence determinants associated with CA-MRSA were common in both wastewater and clinical isolates. PVL genes (*lukF-PV* and *lukS-PV*) were detected in more than one-third of isolates, including over one-quarter of wastewater isolates and 40% of clinical isolates (**Table 2**). More than 95% of isolates carried genes encoding α-, β-, δ-, and γ-hemolysins (**Table S12**). At least one staphylococcal enterotoxin gene was identified in nearly two-thirds of isolates and was more frequent among clinical than wastewater isolates. The most common enterotoxin genes were *sea, sed, sec*, and *sell*. Exfoliative toxin genes (*eta/etb*) were detected only in clinical isolates. The *spa* gene was detected at similar frequencies in clinical and wastewater isolates.

## Discussion

Here, we demonstrated isolation and characterization of MRSA from municipal wastewater and compared results to MRSA from clinics throughout Switzerland. Wastewater isolates shared major STs and *spa* types with clinical isolates and exhibited comparable antimicrobial resistance and virulence profiles. Within the full phylogenetic diversity of the clinical isolates, wastewater isolates were found in only a subset of clusters, with corresponding STs often, though not exclusively, associated with CA- or LA-MRSA. Among the wastewater isolates, six (23%) were phylogenetically distinct from clinical isolates and associated with known livestock-associated MRSA strains. While wastewater captures a mix of MRSA strains, the low overall diversity observed compared to clinical samples highlights its insufficiency as a proxy for clinical epidemiology.

Phylogenetic analysis revealed clustering among wastewater isolates. Most of the wastewater isolates cluster with clinical isolates, except for six ST97 MRSA (23%) that were not related to any clinical isolate in Switzerland. ST97 has well-documented livestock origins: human-adapted ST97 clones are descended from ancestors that circulated in cattle (19). While ST97 has been identified in livestock in Germany and Italy, it has not yet, to our knowledge, been detected in Switzerland (20). The detection of ST97 in wastewater and its phylogenetic distinctness from clinical isolates suggest that wastewater-based surveillance can capture LA-MRSA circulating outside both clinical and livestock-focused surveillance systems, underscoring that wastewater is a nuanced complement to clinical surveillance.

Another cluster of clonal wastewater isolates (ST1482, *spa* type t021, all ≤15 cgSNPs and PVL-negative) was recovered in late June 2023 from multiple WWTPs and is closely related to a PVL-positive ST1482 clinical isolate from Geneva. ST1482 was notably reported in China and Saudi Arabia as community-associated. The temporally coincident but geographically diverse cluster may reflect a period of increased MRSA circulation within Switzerland. Indeed, seasonal variation in MRSA incidence has been reported previously, with peaks occurring in spring or late summer (21,22). Despite these localized clusters, only a small proportion of all isolates belonged to clonal groups. While overall MRSA circulation in Switzerland appears to be driven by multiple co-circulating lineages rather than expansion of a single dominant clone, MRSA from wastewater represents only a small subset of these lineages.

Clinical isolates displayed greater diversity across STs, SCC*mec* types, and *spa* types. This may reflect both methodological and biological factors. Methodologically, the number of wastewater isolates was lower than the number of clinical isolates. Few wastewater isolates were obtained due to the insufficient specificity of the culture method. Of the hundreds of presumptive MRSA colonies recovered on CHROMagar MRSA from wastewater, only 5% were confirmed as MRSA by PCR. Although culture-based recovery of MRSA from wastewater has been reported previously (23–25), selective media developed for clinical specimens often show reduced specificity in environmental samples, where non-target organisms such as methicillin-resistant coagulase-negative staphylococci and *Corynebacterium* spp. can produce similar colony morphologies (26,27). Although MRSA can enter wastewater through activities such as swimming, showering, or handwashing (28,29), MRSA is not primarily an enteric organism (e.g., *Escherichia coli, Enterococcus* spp., or *Klebsiella* spp.) and may therefore enter wastewater less consistently or at lower concentrations (30–32).

Most isolates recovered from both wastewater and clinics carried SCC*mec* type IV, which is historically associated with CA-MRSA. In Switzerland, previous work has shown SCC*mec* type IV increasingly in HA-MRSA (33,34). The distinction between HA- and CA-MRSA lineages is increasingly blurred, as traditional CA-MRSA lineages coexist with or replace traditional HA-MRSA lineages in healthcare settings (8,35,36). PVL gene carriage was also historically associated with CA-MRSA but is increasingly found in HA-MRSA isolates. More than one-third of isolates in this study carried PVL genes, and 90% were notably multidrug-resistant. This combination of virulence and resistance may increase the clinical importance of these lineages.

Importantly, wastewater and clinical isolates should not be interpreted as direct proxies for CA- and HA-MRSA, respectively. Wastewater receives contributions from entire urban catchments, including individuals with community-associated infections, healthcare exposure, long-term care residence, or recent hospitalization, while contemporary clinical MRSA populations increasingly include lineages traditionally associated with community settings.

Several limitations should be considered. First, the number of wastewater isolates was relatively small due to insufficient culture specificity. Given the small sample size, it remains uncertain whether the observed diversity is due to limited sample size or to the limited ability of wastewater to capture the full diversity of clinically relevant MRSA. Alternative selective media, enrichment procedures, or culture-independent methods may improve specificity, increasing the sample size of MRSA isolates. Additionally, although clinical isolates were collected from three large Swiss hospitals representing three different linguistic regions, the sampling framework may be subject to biases related to healthcare utilization, patient selection, diagnostic practices, and temporal variation in case occurrence.

Overall, wastewater-derived MRSA captured a subset of the diversity observed in clinical isolates while sharing major lineages, antimicrobial resistance determinants, and virulence factors. Together, these findings suggest that virulence traits historically associated with CA-MRSA and resistance profiles traditionally associated with HA-MRSA are increasingly co-occurring within circulating MRSA populations, potentially increasing their clinical and epidemiological importance.

## Data availability statement

The sequence data is available in the SRA under BioProject ID PRJNA1497090. The bioinformatic pipeline, implemented using the Snakemake workflow management system, is available at https://github.com/sheenaconforti/ecoli-wgs/. All scripts used for data analysis and figure generation are available at https://github.com/EawagPHH/MRSA_WGS. All sample identification numbers are anonymous, not associated with patient data, and generated for the purpose of this analysis.

## Funding statement

The work was funded through the Swiss National Science Foundation grants 192763 to TRJ and 211422 to SDB, and through a Swiss Federal Office of Public Health grant to Christoph Ort and TRJ.

## Authors’ contributions statement

The author contributions below are in accordance with the CRediT statement. J.K.: conceptualization, methodology, formal analysis, investigation, data curation, visualization, writing (original draft), writing (review and editing), visualization; S.C.: conceptualization, methodology, formal analysis, investigation, data curation, visualization, writing (original draft), writing (review and editing), supervision, visualization; S.D.B.: conceptualization, data curation, writing (review and editing); P.F.: conceptualization, data curation, writing (review and editing); V.G.: conceptualization, data curation, writing (review and editing); T.S.: conceptualization, resources, writing (review and editing); S.H.: conceptualization, data curation, writing (review and editing); T.R.J.: conceptualization, resources, writing (review and editing), supervision, project administration, funding acquisition.

## Acknowledgments

We thank Camille Hablützel, Lea Caduff, and Charlie Gan for their assistance in processing wastewater samples. We thank the Wastewater-based Infectious Disease Surveillance (WISE) group and the PHH group for insightful discussions. We are also grateful to the staff of IDA CDA Lugano (Ticino), ARA Werdhölzli (Zurich), ARA Chur (Graubünden), ARA Sensetal Laupen (Bern), and STEP d’Aïre Genève (Geneva) for providing wastewater samples.

## AI statement

Generative AI tools (ChatGPT, OpenAI) were used solely to support coding for data analysis. No AI tools were used to generate scientific interpretations, text, or conclusions. All code and outputs were reviewed and validated by the authors.

## Notes

### Competing Interest Statement

The authors have declared no competing interest.

### Author Declarations

In January, 2023, Repubblica e Cantone Ticino, Dipartimento della sanita e della socialite, Divisione della salute pubblica, Ufficio di sanita stated that Ethics Committee approval for the project is not needed because the bacterial strains are anonymized, so the research does not fall in the field of application of Human research Act Art. 2 and 3, and does not need approval.

