## Supplemental Material for "Genomic insights into methicillin-resistant *Staphylococcus aureus* from Swiss wastewater and clinical samples"

**Supplementary Material**

1. **PCR conditions to confirm of presumptive MRSA wastewater isolates**

PCR reactions contained 1 µL of DNA and were performed using GoTaq G2 Green Master Mix (Promega) and 10 μM of previously described primers (16SSAIII: 5’-TATAGATGGATCCGCGCT-3’, 16SSAIV: 5’-GATTAGGTACCGTCAAGAT-3’) (1). Cycling conditions consisted of an initial denaturation at 95°C for 1 min, followed by 30 cycles of 95°C for 1 min, 52°C for 1 min, and 72°C for 1 min, with a final extension at 72°C for 5 min. Each run included positive extraction and amplification controls (clinical MRSA) and a negative control (nuclease-free water). PCR products were analyzed by electrophoresis on 2% agarose gels in 1× TAE buffer with a 100-bp ladder (BenchTop, Promega). Isolates producing the expected 273-bp amplicon were confirmed as *S. aureus*.

2. **Bioinformatic analyses of genomic sequences**

All bioinformatic analyses were implemented using the Snakemake workflow management system and are available at <https://github.com/sheenaconforti/ecoli-wgs>.

Illumina reads were trimmed using trimmomatic v0.39 in paired-end mode, removing Illumina Nextera adapters using specific trimming parameters (2:30:10:1) and applying quality-based trimming with a 4bp sliding window and an average quality threshold of 20 (2). Read quality before and after trimming was assessed using FastQC v0.12.1 (3). Screening and filtering for contamination were performed using FastQ Screen v0.16.0 (4) by mapping the trimmed reads against six reference genomes obtained from NCBI: *S. aureus* (ASM1342v1), *Staphylococcus argenteus* (ASM2369v1), *Staphylococcus epidermidis* (ASM609437v1), *Corynebacterium kroppenstedtii* (ASM2314v1), *Enterococcus faecium* (ASM973400v2), *Enterococcus durans* (ASM227793v1), and *Enterococcus lactis* (ASM1934312v1). These species were selected because they have previously been detected or described to grow on CHROMagar MRSA agar plates and could have been mis- or co-cultured with *S. aureus* (5). Filtered reads were *de novo* assembled using SPAdes v4.0.0, without further error correction and set to ’careful’ (6). Quality of the assembled genomes was evaluated with QUAST v5.2.0 (7). Annotation of assembled genomes was performed using Bakta v1.9.4, specifying *Staphylococcus* as genus and *S. aureus* as species (8).

Genome UNClutterer v1.0.6 (GUNC) was used to detect contamination of assembled contigs on a subset of samples (9). Only contigs taxonomically classified as *S. aureus* were kept for the downstream analyses. Decontamination using GUNC was evaluated with QUAST v5.2.0, and the resulting decontaminated genomes were re-annotated using Bakta.

A pangenome was defined using Roary v3.13.0, which generated a core gene alignment for phylogeny reconstruction (10). Core genes were identified as those present in at least 99% of the isolates. Recombinant regions were masked using ClonalFrameML (v.1.13). The resulting masked alignment comprised 116 sequences and 136,081 nucleotide sites, of which 3,079 were parsimony-informative, while 91.1% of sites (123,940 sites) were constant. A maximum likelihood phylogeny was constructed using IQ-Tree v2.3.6 with automatic model selection (11). ModelFinder identified GTR+F+R3 as the best-fitting substitution model based on the Bayesian Information Criterion (BIC) (12). Branch confidence was evaluated using 1000 ultrafast bootstrap replicates (UFBoot) and SH-like approximate likelihood ratio tests (SH-aLRT) (13). The tree was rooted using *S. argenteus* (ASM2369v1) as an outgroup, which is an ancestral *S. aureus* strain recently reclassified as *S. argenteus* (14). The resulting tree was displayed and annotated using iTOL (15). SNP-dist v0.8.2 was used to calculate a core genome single-nucleotide polymorphism (cgSNP) distance matrix between wastewater isolates (16). A maximum of 15 cgSNPs was used to rule out recent transmission of MRSA (within the previous six months) (17).

3. **Isolate characterization**

Isolates were characterized by sequence types (STs), SCC*mec* types, spa types, and the presence of antibiotic resistance genes (ARGs) and virulence factors (VFs). The STs were determined using the seven housekeeping genes of the multilocus sequence typing (MLST) scheme for *S. aureus* v2.23.0 (18). SCC*mec* types were determined using sccmec v1.2.0 (19). Spa types were determined using spaTyper v0.3.3 (20). SpaTyper was run on assembled WGS contigs as well as Sanger DNA sequences. Results from Sanger sequencing were used to complement those from WGS. Abricate v1.0.1 was used to blast the assembled genomes against the Comprehensive Antibiotic Resistance Database (CARD, version of 04.11.2023) to detect ARGs and against the Virulence Factor Database (VFDB, version of 04.11.2023) to detect VFs (21–23). Resistance was defined as the presence of at least one ARG conferring resistance to the relevant antibiotic class. Statistical differences between clinical and wastewater isolates were assessed using the Mann-Whitney U test with ⍺ < 0.05. Statistical analyses were performed in R (v4.4.1) and RStudio (v2024.04.2).

4. **Spa typing by PCR amplification and Sanger sequencing**

To complement WGS-based spa typing and resolve spa types that could not be determined from genome assemblies, the polymorphic region of the S. aureus protein A (spa) gene was amplified and sequenced using primers spa-1113f and spa-1514r as previously described (19). PCR was performed with GoTaq G2 Green Master Mix (Promega) and included a no-template control (nuclease-free water). Thermocycling conditions consisted of initial denaturation at 80°C for 5 min, followed by 35 cycles of 95°C for 45s, 60°C for 1 min, and 72°C for 1 min, and a final extension at 72°C for 10 min. PCR products were analyzed by electrophoresis on 2% agarose gels in 1× TAE buffer with a 100-bp ladder (BenchTop, Promega). The presence of a 400-bp band indicated successful amplification. 20 µL of PCR products were purified using the Wizard SV Gel and PCR Clean-Up System (Promega), following the manufacturer’s instructions and eluting twice in 35 µL of nuclease-free water. 15 µL of purified PCR products were sent to Microsynth (Switzerland) for Sanger sequencing with primer spa-1113f. Chromatograms were manually checked for double peaks using MEGA v12, and the base with the strongest signal was retained.

**Supplementary Figures**

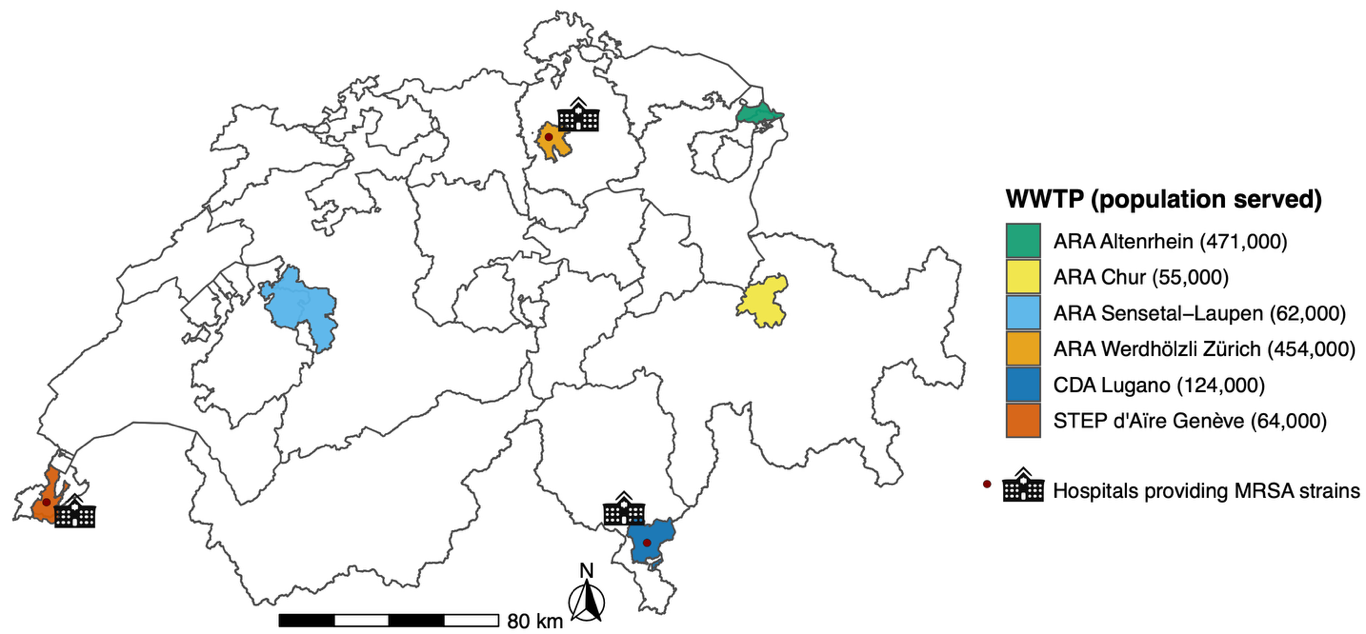

**Figure S1: Geographic distribution of the six investigated wastewater treatment plant (WWTP) catchment areas in Switzerland.** Polygons show each WWTP catchment, colored by plant, with legend entries listing the WWTP name and the estimated population served. Hospital symbols and red circles mark hospitals that provided clinical MRSA isolates, and corresponding placement within the catchment area, including Geneva University Hospitals (HUG, Geneva catchment area), Ente Ospedaliero Cantonale (EOC, samples from hospitals within the Lugano catchment), and University Hospital Zurich (USZ, Zurich catchment area). The figure was generated in R v2025.09.0 and further modified using Inkscape v1.4.2.

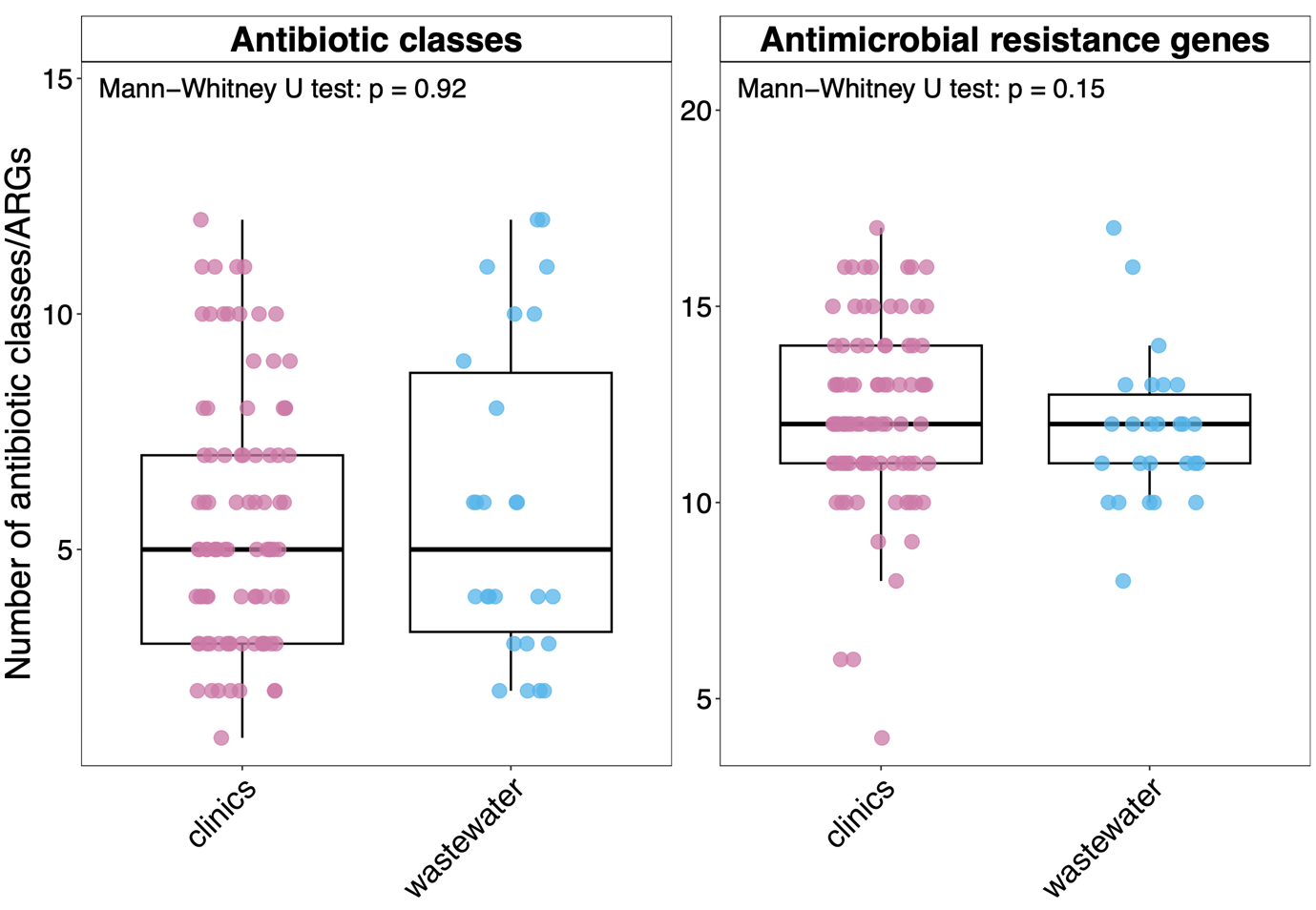

**Figure S2:** Number of antimicrobial resistance genes (ARGs) and antibiotic resistance classes detected in clinical and wastewater MRSA isolates. Resistance to an antibiotic class was defined as the presence of at least one ARG associated with resistance to that class, irrespective of gene expression. ARGs classified as efflux pumps were included in comparisons of ARG counts per isolate but excluded from comparisons of antibiotic resistance classes. Points represent individual isolates; boxes indicate the median and interquartile range. P-values were calculated using two-sided Mann-Whitney U tests.

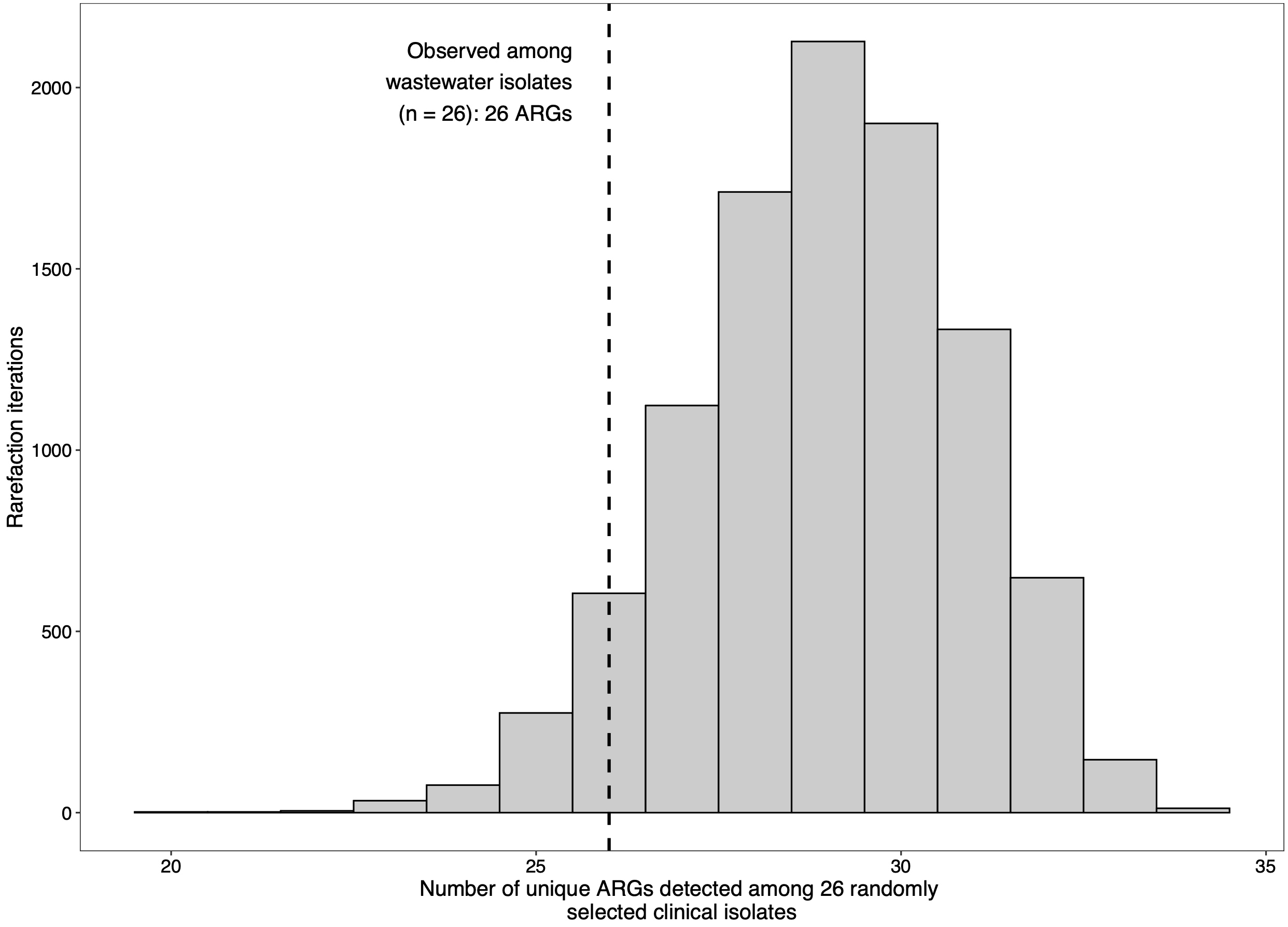

**Figure S3:** Rarefaction analysis of unique antimicrobial resistance genes (ARGs) detected among clinical MRSA isolates. To account for differences in sample size between sources, 26 distinct clinical isolates, matching the number of wastewater isolates, were randomly selected without replacement from the clinical dataset across 10,000 iterations. For each iteration, the number of unique ARGs detected among the selected isolates was recorded. The histogram shows the distribution of the number of unique ARGs across rarefaction iterations. The dashed vertical line indicates the observed number of unique ARGs detected among wastewater isolates (n = 26).

**Supplementary Tables**

**Table S1:** Sampling locations and collection period of methicillin-resistant *Staphylococcus aureus* (MRSA) isolates included in this study. The table reports the number of isolates recovered from each source and location, together with the first and last sampling dates.

| **Source** | **Location** | **No. isolates** | **First sample** | **Last sample** |
| --- | --- | --- | --- | --- |
| clinics | Geneva | 28 | 2022-12-23 | 2023-12-25 |
|  | Ticino | 31 | 2022-12-05 | 2024-02-21 |
|  | Zurich | 30 | 2021-04-25 | 2024-06-14 |
| wastewater | Altenrhein | 5 | 2022-12-13 | 2024-01-08 |
|  | Chur | 8 | 2023-03-07 | 2024-01-07 |
|  | Geneva | 5 | 2023-03-07 | 2024-01-09 |
|  | Laupen | 1 | 2024-01-08 | 2024-01-08 |
|  | Lugano | 6 | 2023-06-27 | 2024-01-08 |
|  | Zurich | 1 | 2023-11-12 | 2023-11-12 |

**Table S2:** Identical MRSA isolates (0 cgSNPs) originating from the same sample and collapsed into a single representative. For each case, the retained isolate is listed alongside the collapsed identical sequences, source compartment, and collection date. This includes multiple clonal isolates from the same wastewater sample.

| **Removed Isolate** | **Retained Isolate** | **Compartment** | **Collection Date** |
| --- | --- | --- | --- |
| 20221213alt3 | 20221213alt4 | wastewater | 13_12_2022 |
| 20230627chu5 | 20230627chu6 | wastewater | 27_06_2023 |
| 20230627chu7 | 20230627chu6 | wastewater | 27_06_2023 |
| 20230627lug6 | 20230627lug7 | wastewater | 27_06_2023 |

**Table S3:** List of antimicrobial resistance genes (ARGs) and associated antibiotic classes as assigned using the Comprehensive Antibiotic Resistance Database (CARD, accessed 25 May 2025) annotations.

| **ARG** | **Antibiotic class** |
| --- | --- |
| AAC(6')-Ie-APH(2'')-Ia | aminoglycoside |
| AAC(6')-Ii | aminoglycoside |
| ANT(4')-Ib | aminoglycoside |
| APH(3')-IIIa | aminoglycoside |
| ErmA | lincosamide, macrolide, streptogramin |
| ErmC | lincosamide, macrolide, streptogramin |
| ErmT | lincosamide, macrolide, streptogramin |
| PC1 beta-lactamase (blaZ) | penam |
| SAT-4 | nucleoside |
| *Staphylococcus aureus* FosB | fosfomycin |
| *Staphylococcus aureus* norA | Efflux pump |
| *Staphylococcus aureus* LmrS | Efflux pump |
| aad(6) | aminoglycoside |
| arlR | Efflux pump |
| arlS | Efflux pump |
| dfrC | diaminopyrimidine |
| dfrG | diaminopyrimidine |
| efmA | Efflux pump |
| fexA | Efflux pump |
| fusB | fusidicacid |
| fusC | fusidicacid |
| lnuA | lincosamide |
| lsaE | lincosamide, macrolide, oxazolidinone,  phenicol, pleuromutilin, streptogramin, tetracycline |
| mecA | penam |
| mecI | penam |
| mecR1 | penam |
| mepA | Efflux pump |
| mepR | Efflux pump |
| efmA | Efflux pump |
| mphC | macrolide |
| msrA | lincosamide, macrolide, oxazolidinone,  phenicol, pleuromutilin, streptogramin, tetracycline |
| msrC | lincosamide, macrolide, oxazolidinone,  phenicol, pleuromutilin, streptogramin, tetracycline |
| mupA | mupirocin |
| tet(38) | tetracycline |
| tet(K) | tetracycline |
| tet(L) | tetracycline |
| tetM | tetracycline |
| mgrA | Efflux pump |

**Table S4**: Number of presumptive MRSA isolates collected at each wastewater treatment plant (WWTP) by sampling date. Values indicate the count of unique presumptive MRSA isolate identifiers detected at each WWTP for each sampling event.

| **Date** | **WWTP**  **Altenrhein** | **WWTP**  **Chur** | **WWTP**  **Geneva** | **WWTP**  **Sensetal-Laupen** | **WWTP**  **Lugano** | **WWTP**  **Zurich** | **Total** |
| --- | --- | --- | --- | --- | --- | --- | --- |
| 12/13/22 | 6 | 7 | 7 | 7 | 7 | 7 | **41** |
| 1/10/23 | 5 | 7 | 5 | 3 | 1 | 0 | **21** |
| 2/7/23 | 0 | 3 | 7 | 6 | 7 | 1 | **24** |
| 3/7/23 | 2 | 5 | 7 | 7 | 7 | 7 | **35** |
| 4/4/23 | 4 | 7 | 7 | 3 | 5 | 6 | **32** |
| 5/2/23 | 7 | 7 | 0 | 7 | 7 | 5 | **33** |
| 5/30/23 | 5 | 2 | 5 | 3 | 2 | 0 | **17** |
| 6/27/23 | 7 | 7 | 7 | 7 | 7 | 6 | **41** |
| 7/23/23 | 0 | 7 | 0 | 0 | 0 | 6 | **13** |
| 7/24/23 | 7 | 0 | 0 | 7 | 6 | 0 | **20** |
| 7/25/23 | 0 | 0 | 7 | 0 | 0 | 0 | **7** |
| 8/20/23 | 0 | 7 | 0 | 0 | 0 | 7 | **14** |
| 8/21/23 | 7 | 0 | 0 | 7 | 7 | 0 | **21** |
| 8/22/23 | 0 | 0 | 7 | 0 | 0 | 0 | **7** |
| 9/17/23 | 0 | 7 | 0 | 0 | 0 | 7 | **14** |
| 9/18/23 | 7 | 0 | 0 | 7 | 7 | 0 | **21** |
| 9/19/23 | 0 | 0 | 7 | 0 | 0 | 0 | **7** |
| 10/17/23 | 0 | 0 | 3 | 0 | 0 | 0 | **3** |
| 10/22/23 | 0 | 7 | 0 | 0 | 0 | 0 | **7** |
| 10/23/23 | 7 | 0 | 0 | 5 | 7 | 0 | **19** |
| 11/12/23 | 0 | 7 | 0 | 0 | 0 | 7 | **14** |
| 11/13/23 | 7 | 0 | 0 | 7 | 7 | 0 | **21** |
| 11/14/23 | 0 | 0 | 5 | 0 | 0 | 0 | **5** |
| 12/10/23 | 0 | 7 | 0 | 0 | 0 | 6 | **13** |
| 12/11/23 | 6 | 0 | 0 | 3 | 7 | 0 | **16** |
| 12/12/23 | 0 | 0 | 7 | 0 | 0 | 0 | **7** |
| 1/7/24 | 0 | 7 | 0 | 0 | 0 | 5 | **12** |
| 1/8/24 | 7 | 0 | 0 | 6 | 7 | 0 | **20** |
| 1/9/24 | 0 | 0 | 7 | 0 | 0 | 0 | **7** |
| **Total** | **84** | **94** | **88** | **85** | **91** | **70** | **512** |

**Table S5**: Comprehensive summary of MRSA sequencing outcomes, quality filtering, and control types by strain.

| **Category** | **N** |
| --- | --- |
| Total unique MRSA isolates | 117 |
| From wastewater | 27 |
| From clinics | 90 |
| Included in final tree (total) | 115 |
| Included in final tree (wastewater) | 26 |
| Included in final tree (clinics) | 89 |
| Excluded from final tree (total) | 2 |
| Exclusion due to contamination | 1 |
| Exclusion due to low coverage | 1 |
| Exclusion due to low assembly quality | 0 |
| AE negative control samples | 2 |
| *E. coli* strain1 | 1 |
| *E. coli* strain2 | 1 |

**Table S6**: Distribution of methicillin-resistant *Staphylococcus aureus* (MRSA) isolates by sequence type (ST), identified by the analysis of seven housekeeping genes of the multilocus sequence typing (MLST)20& Achtman scheme using MLST v.2.16.1 (<https://github.com/tseemann/mlst>). The table includes the number and percentage of isolates for each ST. Sequence types not determined are assigned na.

| **Sequence Type** | **Number (percentage %) of total isolates** | **Number (percentage %) of clinical isolates** | **Number (percentage %) of wastewater isolates** |
| --- | --- | --- | --- |
| 22 | 20 (17.4%) | 17 (19.1%) | 3 (11.5%) |
| 5 | 15 (13%) | 15 (16.9%) | 0 (0%) |
| na | 15 (13%) | 10 (11.2%) | 5 (19.2%) |
| 8 | 9 (7.8%) | 9 (10.1%) | 0 (0%) |
| 1482 | 7 (6.1%) | 1 (1.1%) | 6 (23.1%) |
| 6 | 6 (5.2%) | 2 (2.2%) | 4 (15.4%) |
| 1 | 5 (4.3%) | 5 (5.6%) | 0 (0%) |
| 97 | 5 (4.3%) | 0 (0%) | 5 (19.2%) |
| 1153 | 4 (3.5%) | 4 (4.5%) | 0 (0%) |
| 152 | 4 (3.5%) | 3 (3.4%) | 1 (3.8%) |
| 88 | 4 (3.5%) | 2 (2.2%) | 2 (7.7%) |
| 225 | 3 (2.6%) | 3 (3.4%) | 0 (0%) |
| 30 | 3 (2.6%) | 3 (3.4%) | 0 (0%) |
| 398 | 2 (1.7%) | 2 (2.2%) | 0 (0%) |
| 45 | 2 (1.7%) | 2 (2.2%) | 0 (0%) |
| 672 | 2 (1.7%) | 2 (2.2%) | 0 (0%) |
| 105 | 1 (0.9%) | 1 (1.1%) | 0 (0%) |
| 1232 | 1 (0.9%) | 1 (1.1%) | 0 (0%) |
| 1535 | 1 (0.9%) | 1 (1.1%) | 0 (0%) |
| 1635 | 1 (0.9%) | 1 (1.1%) | 0 (0%) |
| 5969 | 1 (0.9%) | 1 (1.1%) | 0 (0%) |
| 6659 | 1 (0.9%) | 1 (1.1%) | 0 (0%) |
| 6692 | 1 (0.9%) | 1 (1.1%) | 0 (0%) |
| 7 | 1 (0.9%) | 1 (1.1%) | 0 (0%) |
| 80 | 1 (0.9%) | 1 (1.1%) | 0 (0%) |

**Table S7:** Distribution of methicillin-resistant *Staphylococcus aureus* (MRSA) isolates by SCC*mec* type, stratified by source. SCC*mec* types not determined are assigned na.

| **SCC mec** | **Number (percentage %) of total isolates** | **Number (percentage %) of clinical isolates** | **Number (percentage %) of wastewater isolates** |
| --- | --- | --- | --- |
| **IIa** | 4 (3.5%) | 4 (4.5%) | 0 (0%) |
| **IId** | 3 (2.6%) | 3 (3.4%) | 0 (0%) |
| **IVa** | 47 (40.9%) | 27 (30.3%) | 20 (76.9%) |
| **IVb** | 24 (20.9%) | 18 (20.2%) | 6 (23.1%) |
| **IVc** | 1 (0.9%) | 1 (1.1%) | 0 (0%) |
| **IVg** | 1 (0.9%) | 1 (1.1%) | 0 (0%) |
| **IVj** | 10 (8.7%) | 10 (11.2%) | 0 (0%) |
| **V** | 8 (7%) | 8 (9%) | 0 (0%) |
| **VI** | 5 (4.3%) | 5 (5.6%) | 0 (0%) |
| **Va** | 4 (3.5%) | 4 (4.5%) | 0 (0%) |
| **Vc** | 6 (5.2%) | 6 (6.7%) | 0 (0%) |
| **na** | 2 (1.7%) | 2 (2.2%) | 0 (0%) |

**Table S8**: Distribution of methicillin-resistant *Staphylococcus aureus* (MRSA) isolates by spa type, stratified by source. Spa types not determined by either method (WGS or Sanger) are assigned na.

| **spa** | **Number (percentage %) of total isolates** | **Number (percentage %) of clinical isolates** | **Number (percentage %) of wastewater isolates** |
| --- | --- | --- | --- |
| **na** | 14 (12.2%) | 13 (14.6%) | 1 (3.8%) |
| **t002** | 9 (7.8%) | 9 (10.1%) | 0 (0%) |
| **t003** | 2 (1.7%) | 2 (2.2%) | 0 (0%) |
| **t005** | 4 (3.5%) | 4 (4.5%) | 0 (0%) |
| **t008** | 4 (3.5%) | 4 (4.5%) | 0 (0%) |
| **t019** | 2 (1.7%) | 2 (2.2%) | 0 (0%) |
| **t021** | 7 (6.1%) | 1 (1.1%) | 6 (23.1%) |
| **t022** | 1 (0.9%) | 1 (1.1%) | 0 (0%) |
| **t026** | 2 (1.7%) | 2 (2.2%) | 0 (0%) |
| **t030** | 1 (0.9%) | 1 (1.1%) | 0 (0%) |
| **t032** | 1 (0.9%) | 1 (1.1%) | 0 (0%) |
| **t034** | 1 (0.9%) | 1 (1.1%) | 0 (0%) |
| **t044** | 1 (0.9%) | 1 (1.1%) | 0 (0%) |
| **t045** | 1 (0.9%) | 1 (1.1%) | 0 (0%) |
| **t084** | 1 (0.9%) | 1 (1.1%) | 0 (0%) |
| **t091** | 1 (0.9%) | 1 (1.1%) | 0 (0%) |
| **t105** | 1 (0.9%) | 1 (1.1%) | 0 (0%) |
| **t10682** | 1 (0.9%) | 1 (1.1%) | 0 (0%) |
| **t112** | 1 (0.9%) | 1 (1.1%) | 0 (0%) |
| **t127** | 4 (3.5%) | 4 (4.5%) | 0 (0%) |
| **t1437** | 1 (0.9%) | 1 (1.1%) | 0 (0%) |
| **t1476** | 2 (1.7%) | 2 (2.2%) | 0 (0%) |
| **t177** | 1 (0.9%) | 1 (1.1%) | 0 (0%) |
| **t1815** | 1 (0.9%) | 1 (1.1%) | 0 (0%) |
| **t223** | 6 (5.2%) | 3 (3.4%) | 3 (11.5%) |
| **t2453** | 1 (0.9%) | 0 (0%) | 1 (3.8%) |
| **t304** | 7 (6.1%) | 2 (2.2%) | 5 (19.2%) |
| **t309** | 1 (0.9%) | 1 (1.1%) | 0 (0%) |
| **t311** | 1 (0.9%) | 1 (1.1%) | 0 (0%) |
| **t355** | 3 (2.6%) | 2 (2.2%) | 1 (3.8%) |
| **t359** | 2 (1.7%) | 0 (0%) | 2 (7.7%) |
| **t3614** | 1 (0.9%) | 1 (1.1%) | 0 (0%) |
| **t3841** | 2 (1.7%) | 2 (2.2%) | 0 (0%) |
| **t4103** | 1 (0.9%) | 1 (1.1%) | 0 (0%) |
| **t442** | 1 (0.9%) | 1 (1.1%) | 0 (0%) |
| **t454** | 1 (0.9%) | 1 (1.1%) | 0 (0%) |
| **t504** | 2 (1.7%) | 2 (2.2%) | 0 (0%) |
| **t517** | 8 (7%) | 2 (2.2%) | 6 (23.1%) |
| **t535** | 1 (0.9%) | 1 (1.1%) | 0 (0%) |
| **t571** | 1 (0.9%) | 1 (1.1%) | 0 (0%) |
| **t586** | 1 (0.9%) | 1 (1.1%) | 0 (0%) |
| **t605** | 1 (0.9%) | 0 (0%) | 1 (3.8%) |
| **t6172** | 1 (0.9%) | 1 (1.1%) | 0 (0%) |
| **t648** | 1 (0.9%) | 1 (1.1%) | 0 (0%) |
| **t685** | 1 (0.9%) | 1 (1.1%) | 0 (0%) |
| **t688** | 3 (2.6%) | 3 (3.4%) | 0 (0%) |
| **t6978** | 1 (0.9%) | 1 (1.1%) | 0 (0%) |
| **t7418** | 1 (0.9%) | 1 (1.1%) | 0 (0%) |
| **t903** | 2 (1.7%) | 2 (2.2%) | 0 (0%) |

**Table S9:** Distribution of 63 unique ST-SCCmec-spa combinations among MRSA isolates. Isolates with undefined ST, SCC*mec*, or spa type assignments were excluded.

| **ST-SCC*mec*-spa combination** | **Total isolates, n (%)** | **Clinical isolates, n (%)** | **Wastewater isolates, n (%)** |
| --- | --- | --- | --- |
| 1482-IVa-t021 | 7 (7.6%) | 1 (1.4%) | 6 (28.6%) |
| 5-IVb-t002 | 6 (6.5%) | 6 (8.5%) | 0 (0%) |
| 6-IVa-t304 | 5 (5.4%) | 2 (2.8%) | 3 (14.3%) |
| 22-IVa-t005 | 4 (4.3%) | 4 (5.6%) | 0 (0%) |
| 22-IVb-t223 | 3 (3.3%) | 1 (1.4%) | 2 (9.5%) |
| 88-IVa-t517 | 3 (3.3%) | 1 (1.4%) | 2 (9.5%) |
| 1153-IVb-t504 | 2 (2.2%) | 2 (2.8%) | 0 (0%) |
| 152-IVa-t355 | 2 (2.2%) | 1 (1.4%) | 1 (4.8%) |
| 225-IIa-t003 | 2 (2.2%) | 2 (2.8%) | 0 (0%) |
| 45-IVa-t026 | 2 (2.2%) | 2 (2.8%) | 0 (0%) |
| 5-VI-t688 | 2 (2.2%) | 2 (2.8%) | 0 (0%) |
| 8-IVa-t008 | 2 (2.2%) | 2 (2.8%) | 0 (0%) |
| 97-IVa-t359 | 2 (2.2%) | 0 (0%) | 2 (9.5%) |
| 1-IVb-t127 | 1 (1.1%) | 1 (1.4%) | 0 (0%) |
| 1-IVb-t177 | 1 (1.1%) | 1 (1.4%) | 0 (0%) |
| 1-V-t127 | 1 (1.1%) | 1 (1.4%) | 0 (0%) |
| 1-VI-t127 | 1 (1.1%) | 1 (1.4%) | 0 (0%) |
| 1-Va-t127 | 1 (1.1%) | 1 (1.4%) | 0 (0%) |
| 105-IIa-t10682 | 1 (1.1%) | 1 (1.4%) | 0 (0%) |
| 1153-IVa-t903 | 1 (1.1%) | 1 (1.4%) | 0 (0%) |
| 1153-Va-t903 | 1 (1.1%) | 1 (1.4%) | 0 (0%) |
| 1232-V-t034 | 1 (1.1%) | 1 (1.4%) | 0 (0%) |
| 152-IVb-t355 | 1 (1.1%) | 1 (1.4%) | 0 (0%) |
| 152-Va-t454 | 1 (1.1%) | 1 (1.4%) | 0 (0%) |
| 1535-V-t084 | 1 (1.1%) | 1 (1.4%) | 0 (0%) |
| 1635-IVa-t002 | 1 (1.1%) | 1 (1.4%) | 0 (0%) |
| 22-IVa-t112 | 1 (1.1%) | 1 (1.4%) | 0 (0%) |
| 22-IVa-t1476 | 1 (1.1%) | 1 (1.4%) | 0 (0%) |
| 22-IVa-t223 | 1 (1.1%) | 0 (0%) | 1 (4.8%) |
| 22-IVa-t309 | 1 (1.1%) | 1 (1.4%) | 0 (0%) |
| 22-IVa-t6978 | 1 (1.1%) | 1 (1.4%) | 0 (0%) |
| 22-IVj-t022 | 1 (1.1%) | 1 (1.4%) | 0 (0%) |
| 22-IVj-t032 | 1 (1.1%) | 1 (1.4%) | 0 (0%) |
| 22-IVj-t1437 | 1 (1.1%) | 1 (1.4%) | 0 (0%) |
| 22-IVj-t7418 | 1 (1.1%) | 1 (1.4%) | 0 (0%) |
| 225-IIa-t586 | 1 (1.1%) | 1 (1.4%) | 0 (0%) |
| 30-IVa-t019 | 1 (1.1%) | 1 (1.4%) | 0 (0%) |
| 30-IVa-t030 | 1 (1.1%) | 1 (1.4%) | 0 (0%) |
| 30-IVj-t019 | 1 (1.1%) | 1 (1.4%) | 0 (0%) |
| 398-IVc-t517 | 1 (1.1%) | 1 (1.4%) | 0 (0%) |
| 5-IId-t002 | 1 (1.1%) | 1 (1.4%) | 0 (0%) |
| 5-IVa-t105 | 1 (1.1%) | 1 (1.4%) | 0 (0%) |
| 5-IVb-t045 | 1 (1.1%) | 1 (1.4%) | 0 (0%) |
| 5-V-t535 | 1 (1.1%) | 1 (1.4%) | 0 (0%) |
| 5-VI-t1815 | 1 (1.1%) | 1 (1.4%) | 0 (0%) |
| 5-Va-t442 | 1 (1.1%) | 1 (1.4%) | 0 (0%) |
| 5969-IVb-t008 | 1 (1.1%) | 1 (1.4%) | 0 (0%) |
| 6-IVa-t517 | 1 (1.1%) | 0 (0%) | 1 (4.8%) |
| 6659-Vc-t223 | 1 (1.1%) | 1 (1.4%) | 0 (0%) |
| 6692-IVa-t688 | 1 (1.1%) | 1 (1.4%) | 0 (0%) |
| 672-V-t3841 | 1 (1.1%) | 1 (1.4%) | 0 (0%) |
| 672-Vc-t3841 | 1 (1.1%) | 1 (1.4%) | 0 (0%) |
| 8-IVa-t6172 | 1 (1.1%) | 1 (1.4%) | 0 (0%) |
| 8-IVa-t648 | 1 (1.1%) | 1 (1.4%) | 0 (0%) |
| 8-IVb-t008 | 1 (1.1%) | 1 (1.4%) | 0 (0%) |
| 8-IVb-t3614 | 1 (1.1%) | 1 (1.4%) | 0 (0%) |
| 8-V-t1476 | 1 (1.1%) | 1 (1.4%) | 0 (0%) |
| 8-V-t223 | 1 (1.1%) | 1 (1.4%) | 0 (0%) |
| 80-IVb-t044 | 1 (1.1%) | 1 (1.4%) | 0 (0%) |
| 88-IVa-t4103 | 1 (1.1%) | 1 (1.4%) | 0 (0%) |
| 97-IVa-t304 | 1 (1.1%) | 0 (0%) | 1 (4.8%) |
| 97-IVb-t2453 | 1 (1.1%) | 0 (0%) | 1 (4.8%) |
| 97-IVb-t605 | 1 (1.1%) | 0 (0%) | 1 (4.8%) |

**Table S10:** Distribution of MRSA isolates by number of antimicrobial resistance classes inferred from ARGs detected in the sequencing data. Values indicate the number and percentage of isolates carrying ARGs associated with each number of resistance classes, based on Comprehensive Antibiotic Resistance Database (CARD) annotations. Multidrug resistance (MDR) was defined as the presence of ARGs associated with resistance to three or more antimicrobial classes. The “efflux pumps” category was not considered an antimicrobial resistance class.

| **Number of antibiotic classes** | **Overall isolates, n (%)** | **Clinical isolates, n (%)** | **Wastewater isolates, n (%)** |
| --- | --- | --- | --- |
| 1 | 1 (0.9%) | 1 (1.1%) | 0 (0%) |
| 2 | 11 (9.6%) | 7 (7.9%) | 4 (15.4%) |
| 3 | 19 (16.5%) | 16 (18%) | 3 (11.5%) |
| 4 | 17 (14.8%) | 11 (12.4%) | 6 (23.1%) |
| 5 | 15 (13%) | 15 (16.9%) | 0 (0%) |
| 6 | 14 (12.2%) | 9 (10.1%) | 5 (19.2%) |
| 7 | 9 (7.8%) | 9 (10.1%) | 0 (0%) |
| 8 | 7 (6.1%) | 6 (6.7%) | 1 (3.8%) |
| 9 | 4 (3.5%) | 3 (3.4%) | 1 (3.8%) |
| 10 | 9 (7.8%) | 7 (7.9%) | 2 (7.7%) |
| 11 | 6 (5.2%) | 4 (4.5%) | 2 (7.7%) |
| 12 | 3 (2.6%) | 1 (1.1%) | 2 (7.7%) |
| **MDR ≥ 3** | **103 (89.6%)** | **81 (91%)** | **22 (84.6%)** |

**Table S11:** Distribution of antimicrobial resistance genes (ARGs) among MRSA isolates recovered from clinical and wastewater sources. Values indicate the number and percentage of isolates carrying each ARG, overall and stratified by source. ARGs were identified using the Comprehensive Antibiotic Resistance Database (CARD, accessed 25 May 2025).

| **Gene** | **Overall isolates, n (%)** | **Clinical isolates, n (%)** | **Wastewater isolates, n (%)** |
| --- | --- | --- | --- |
| *mepR* | 111 (96.5%) | 87 (97.8%) | 24 (92.3%) |
| *mgrA* | 111 (96.5%) | 86 (96.6%) | 25 (96.2%) |
| *arlR* | 109 (94.8%) | 84 (94.4%) | 25 (96.2%) |
| *mecA* | 109 (94.8%) | 84 (94.4%) | 25 (96.2%) |
| *mepA* | 109 (94.8%) | 85 (95.5%) | 24 (92.3%) |
| *S. aureus norA* | 108 (93.9%) | 82 (92.1%) | 26 (100%) |
| *S. aureus LmrS* | 107 (93%) | 84 (94.4%) | 23 (88.5%) |
| *tet*(38) | 107 (93%) | 82 (92.1%) | 25 (96.2%) |
| *arlS* | 104 (90.4%) | 81 (91%) | 23 (88.5%) |
| PC1 **β** -lactamase_(*blaZ*) | 97 (84.3%) | 79 (88.8%) | 18 (69.2%) |
| *S. aureus FosB* | 65 (56.5%) | 49 (55.1%) | 16 (61.5%) |
| *ErmC* | 33 (28.7%) | 23 (25.8%) | 10 (38.5%) |
| AAC(6')-Ie-APH(2'')-Ia | 28 (24.3%) | 24 (27%) | 4 (15.4%) |
| *dfrC* | 27 (23.5%) | 20 (22.5%) | 7 (26.9%) |
| *tet*(K) | 24 (20.9%) | 20 (22.5%) | 4 (15.4%) |
| *fusC* | 23 (20%) | 23 (25.8%) | 0 (0%) |
| *msrA* | 21 (18.3%) | 15 (16.9%) | 6 (23.1%) |
| *dfrG* | 20 (17.4%) | 14 (15.7%) | 6 (23.1%) |
| *fusB* | 14 (12.2%) | 8 (9%) | 6 (23.1%) |
| APH(3')-IIIa | 11 (9.6%) | 10 (11.2%) | 1 (3.8%) |
| *mphC* | 11 (9.6%) | 8 (9%) | 3 (11.5%) |
| SAT-4 | 9 (7.8%) | 8 (9%) | 1 (3.8%) |
| *ErmA* | 8 (7%) | 8 (9%) | 0 (0%) |
| ANT(4')-Ib | 5 (4.3%) | 4 (4.5%) | 1 (3.8%) |
| *lnuA* | 5 (4.3%) | 4 (4.5%) | 1 (3.8%) |
| *fexA* | 4 (3.5%) | 4 (4.5%) | 0 (0%) |
| *mecI* | 4 (3.5%) | 4 (4.5%) | 0 (0%) |
| *mecR1* | 4 (3.5%) | 4 (4.5%) | 0 (0%) |
| *tetM* | 4 (3.5%) | 4 (4.5%) | 0 (0%) |
| *aad*(6) | 2 (1.7%) | 2 (2.2%) | 0 (0%) |
| AAC(6')-Ii | 1 (0.9%) | 0 (0%) | 1 (3.8%) |
| *ErmT* | 1 (0.9%) | 1 (1.1%) | 0 (0%) |
| *efmA* | 1 (0.9%) | 0 (0%) | 1 (3.8%) |
| *lsaE* | 1 (0.9%) | 1 (1.1%) | 0 (0%) |
| *msrC* | 1 (0.9%) | 0 (0%) | 1 (3.8%) |
| mupA | 1 (0.9%) | 1 (1.1%) | 0 (0%) |
| tet(L) | 1 (0.9%) | 1 (1.1%) | 0 (0%) |

**Table S12:** Distribution of virulence factor genes among MRSA isolates. Values indicate the number and percentage of isolates carrying each gene and are reported as n/N (%), where N is the number of isolates within the corresponding category. Genes were considered present when sequence identity was ≥80%. The row lukF-PV + lukS-PV indicates isolates carrying both PVL subunit genes. Virulence factor genes not assigned to the predefined categories were grouped as other virulence factors.

| **VF Category** | **Gene** | **Overall isolates, n/N (%)** | **Clinical isolates, n/N (%)** | **Wastewater isolates, n/N (%)** |
| --- | --- | --- | --- | --- |
| Panton-Valentine leukocidin | lukF-PV + lukS-PV | 43/115 (37.4%) | 36/89 (40.4%) | 7/26 (26.9%) |
|  | lukF-PV | 85/115 (73.9%) | 69/89 (77.5%) | 16/26 (61.5%) |
|  | lukS-PV | 43/115 (37.4%) | 36/89 (40.4%) | 7/26 (26.9%) |
| Staphylococcal enterotoxins | sea | 49/115 (42.6%) | 37/89 (41.6%) | 12/26 (46.2%) |
|  | seb | 1/115 (0.9%) | 1/89 (1.1%) | 0/26 (0%) |
|  | sec | 17/115 (14.8%) | 17/89 (19.1%) | 0/26 (0%) |
|  | sed | 28/115 (24.3%) | 23/89 (25.8%) | 5/26 (19.2%) |
|  | seh | 10/115 (8.7%) | 9/89 (10.1%) | 1/26 (3.8%) |
|  | selk | 12/115 (10.4%) | 8/89 (9%) | 4/26 (15.4%) |
|  | sell | 17/115 (14.8%) | 17/89 (19.1%) | 0/26 (0%) |
|  | selq | 12/115 (10.4%) | 8/89 (9%) | 4/26 (15.4%) |
| Hemolysins | hlb | 110/115 (95.7%) | 85/89 (95.5%) | 25/26 (96.2%) |
|  | hld | 111/115 (96.5%) | 85/89 (95.5%) | 26/26 (100%) |
|  | hlgA | 111/115 (96.5%) | 85/89 (95.5%) | 26/26 (100%) |
|  | hlgB | 109/115 (94.8%) | 84/89 (94.4%) | 25/26 (96.2%) |
|  | hlgC | 110/115 (95.7%) | 86/89 (96.6%) | 24/26 (92.3%) |
|  | hly/hla | 110/115 (95.7%) | 85/89 (95.5%) | 25/26 (96.2%) |
| Exfoliative toxins | eta | 4/115 (3.5%) | 4/89 (4.5%) | 0/26 (0%) |
|  | etb | 2/115 (1.7%) | 2/89 (2.2%) | 0/26 (0%) |
| Staphylococcal protein A | spa | 13/115 (11.3%) | 10/89 (11.2%) | 3/26 (11.5%) |
| Other virulence factors | acm | 1/115 (0.9%) | 0/89 (0%) | 1/26 (3.8%) |
|  | adsA | 101/115 (87.8%) | 80/89 (89.9%) | 21/26 (80.8%) |
|  | aur | 108/115 (93.9%) | 86/89 (96.6%) | 22/26 (84.6%) |
|  | cap8A | 111/115 (96.5%) | 85/89 (95.5%) | 26/26 (100%) |
|  | cap8B | 111/115 (96.5%) | 87/89 (97.8%) | 24/26 (92.3%) |
|  | cap8C | 109/115 (94.8%) | 85/89 (95.5%) | 24/26 (92.3%) |
|  | cap8D | 103/115 (89.6%) | 81/89 (91%) | 22/26 (84.6%) |
|  | cap8E | 109/115 (94.8%) | 84/89 (94.4%) | 25/26 (96.2%) |
|  | cap8F | 104/115 (90.4%) | 81/89 (91%) | 23/26 (88.5%) |
|  | cap8G | 106/115 (92.2%) | 84/89 (94.4%) | 22/26 (84.6%) |
|  | cap8H | 42/115 (36.5%) | 24/89 (27%) | 18/26 (69.2%) |
|  | cap8I | 45/115 (39.1%) | 27/89 (30.3%) | 18/26 (69.2%) |
|  | cap8J | 47/115 (40.9%) | 29/89 (32.6%) | 18/26 (69.2%) |
|  | cap8K | 45/115 (39.1%) | 27/89 (30.3%) | 18/26 (69.2%) |
|  | cap8L | 106/115 (92.2%) | 83/89 (93.3%) | 23/26 (88.5%) |
|  | cap8M | 111/115 (96.5%) | 86/89 (96.6%) | 25/26 (96.2%) |
|  | cap8N | 109/115 (94.8%) | 85/89 (95.5%) | 24/26 (92.3%) |
|  | cap8O | 106/115 (92.2%) | 83/89 (93.3%) | 23/26 (88.5%) |
|  | cap8P | 107/115 (93%) | 84/89 (94.4%) | 23/26 (88.5%) |
|  | chp | 59/115 (51.3%) | 44/89 (49.4%) | 15/26 (57.7%) |
|  | cna | 1/115 (0.9%) | 0/89 (0%) | 1/26 (3.8%) |
|  | coa | 56/115 (48.7%) | 51/89 (57.3%) | 5/26 (19.2%) |
|  | ebp | 100/115 (87%) | 76/89 (85.4%) | 24/26 (92.3%) |
|  | esaA | 104/115 (90.4%) | 81/89 (91%) | 23/26 (88.5%) |
|  | esaB | 111/115 (96.5%) | 87/89 (97.8%) | 24/26 (92.3%) |
|  | esaC | 83/115 (72.2%) | 66/89 (74.2%) | 17/26 (65.4%) |
|  | essA | 111/115 (96.5%) | 85/89 (95.5%) | 26/26 (100%) |
|  | essB | 108/115 (93.9%) | 86/89 (96.6%) | 22/26 (84.6%) |
|  | essC | 66/115 (57.4%) | 54/89 (60.7%) | 12/26 (46.2%) |
|  | esxA | 114/115 (99.1%) | 88/89 (98.9%) | 26/26 (100%) |
|  | esxB | 79/115 (68.7%) | 65/89 (73%) | 14/26 (53.8%) |
|  | fnbA | 3/115 (2.6%) | 2/89 (2.2%) | 1/26 (3.8%) |
|  | fnbB | 1/115 (0.9%) | 1/89 (1.1%) | 0/26 (0%) |
|  | geh | 103/115 (89.6%) | 82/89 (92.1%) | 21/26 (80.8%) |
|  | hysA | 100/115 (87%) | 81/89 (91%) | 19/26 (73.1%) |
|  | icaA | 110/115 (95.7%) | 86/89 (96.6%) | 24/26 (92.3%) |
|  | icaB | 113/115 (98.3%) | 87/89 (97.8%) | 26/26 (100%) |
|  | icaC | 103/115 (89.6%) | 84/89 (94.4%) | 19/26 (73.1%) |
|  | icaD | 110/115 (95.7%) | 86/89 (96.6%) | 24/26 (92.3%) |
|  | icaR | 109/115 (94.8%) | 85/89 (95.5%) | 24/26 (92.3%) |
|  | isdA | 111/115 (96.5%) | 88/89 (98.9%) | 23/26 (88.5%) |
|  | isdB | 108/115 (93.9%) | 84/89 (94.4%) | 24/26 (92.3%) |
|  | isdC | 112/115 (97.4%) | 87/89 (97.8%) | 25/26 (96.2%) |
|  | isdD | 110/115 (95.7%) | 86/89 (96.6%) | 24/26 (92.3%) |
|  | isdE | 110/115 (95.7%) | 87/89 (97.8%) | 23/26 (88.5%) |
|  | isdF | 109/115 (94.8%) | 86/89 (96.6%) | 23/26 (88.5%) |
|  | isdG | 115/115 (100%) | 89/89 (100%) | 26/26 (100%) |
|  | lip | 106/115 (92.2%) | 83/89 (93.3%) | 23/26 (88.5%) |
|  | map | 86/115 (74.8%) | 67/89 (75.3%) | 19/26 (73.1%) |
|  | sak | 99/115 (86.1%) | 74/89 (83.1%) | 25/26 (96.2%) |
|  | sbi | 102/115 (88.7%) | 79/89 (88.8%) | 23/26 (88.5%) |
|  | scm | 1/115 (0.9%) | 0/89 (0%) | 1/26 (3.8%) |
|  | scn | 103/115 (89.6%) | 78/89 (87.6%) | 25/26 (96.2%) |
|  | sdrD | 19/115 (16.5%) | 12/89 (13.5%) | 7/26 (26.9%) |
|  | sdrE | 33/115 (28.7%) | 30/89 (33.7%) | 3/26 (11.5%) |
|  | sgrA | 2/115 (1.7%) | 0/89 (0%) | 2/26 (7.7%) |
|  | srtB | 111/115 (96.5%) | 86/89 (96.6%) | 25/26 (96.2%) |
|  | sspA | 106/115 (92.2%) | 82/89 (92.1%) | 24/26 (92.3%) |
|  | sspB | 107/115 (93%) | 85/89 (95.5%) | 22/26 (84.6%) |
|  | sspC | 112/115 (97.4%) | 86/89 (96.6%) | 26/26 (100%) |
|  | tsst-1 | 25/115 (21.7%) | 22/89 (24.7%) | 3/26 (11.5%) |
|  | vWbp | 48/115 (41.7%) | 33/89 (37.1%) | 15/26 (57.7%) |
