## Supplementary figures and images for "Genomic insights into methicillin-resistant *Staphylococcus aureus* from Swiss wastewater and clinical samples"

### Supplemental Figure S1

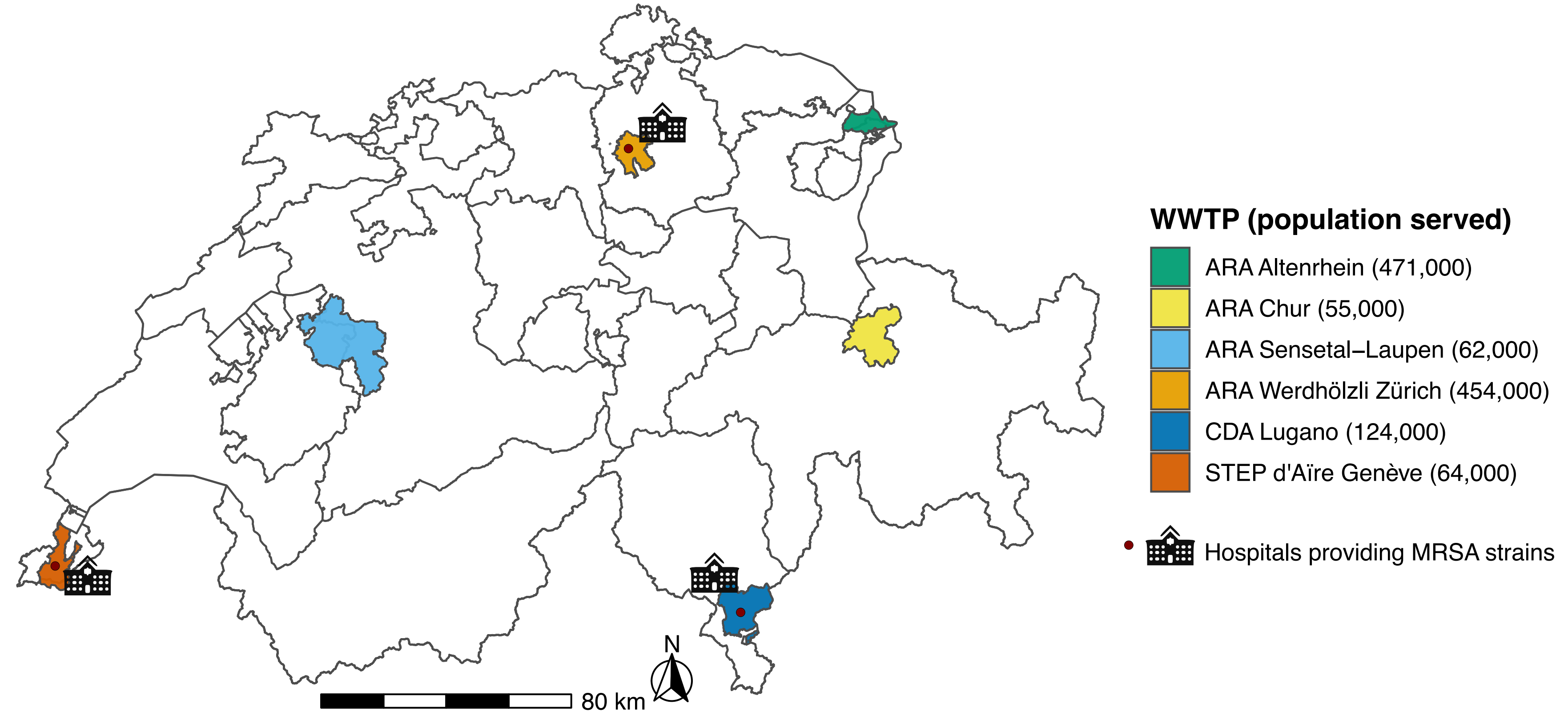

### Supplemental Figure S2

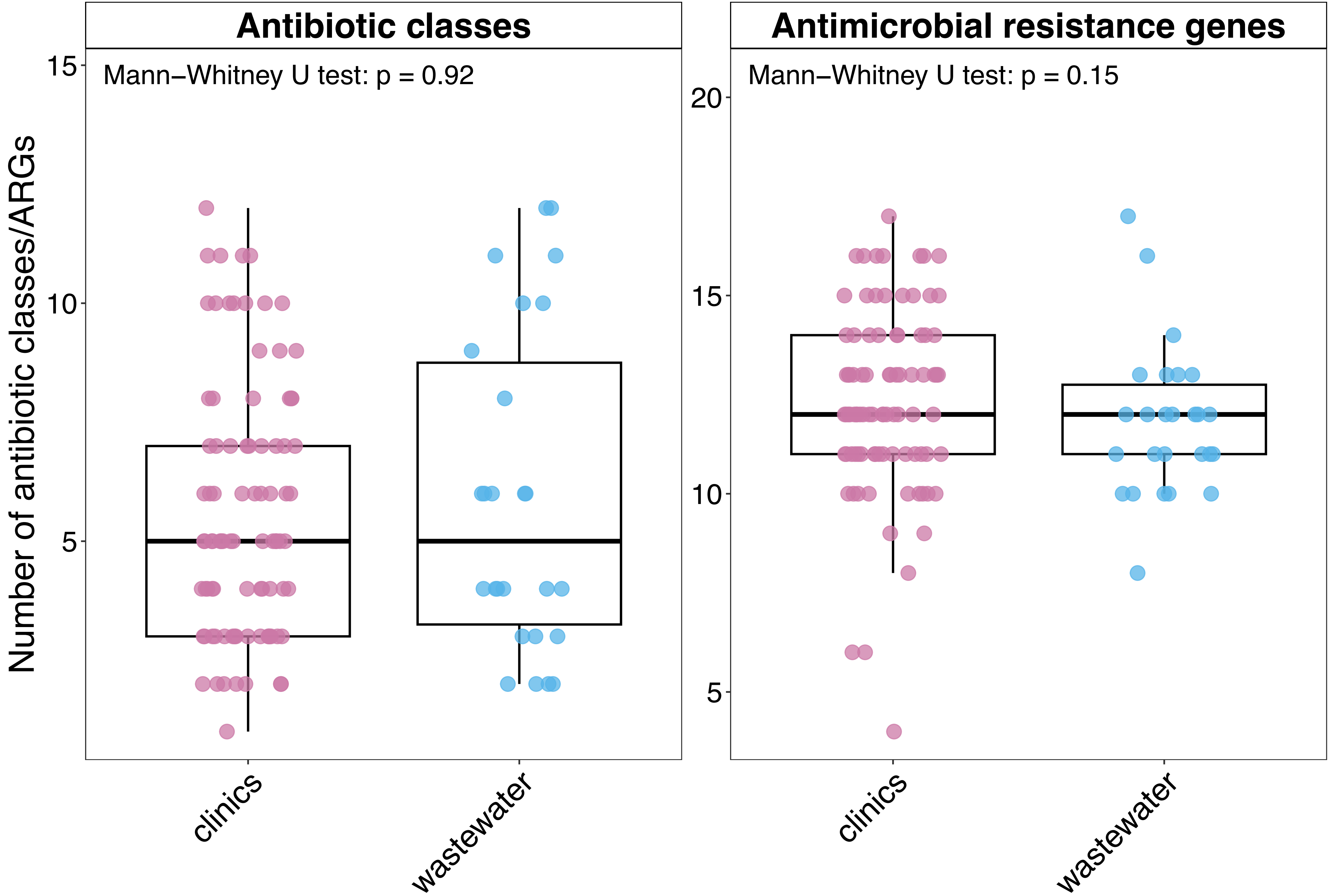

### Supplemental Figure S3

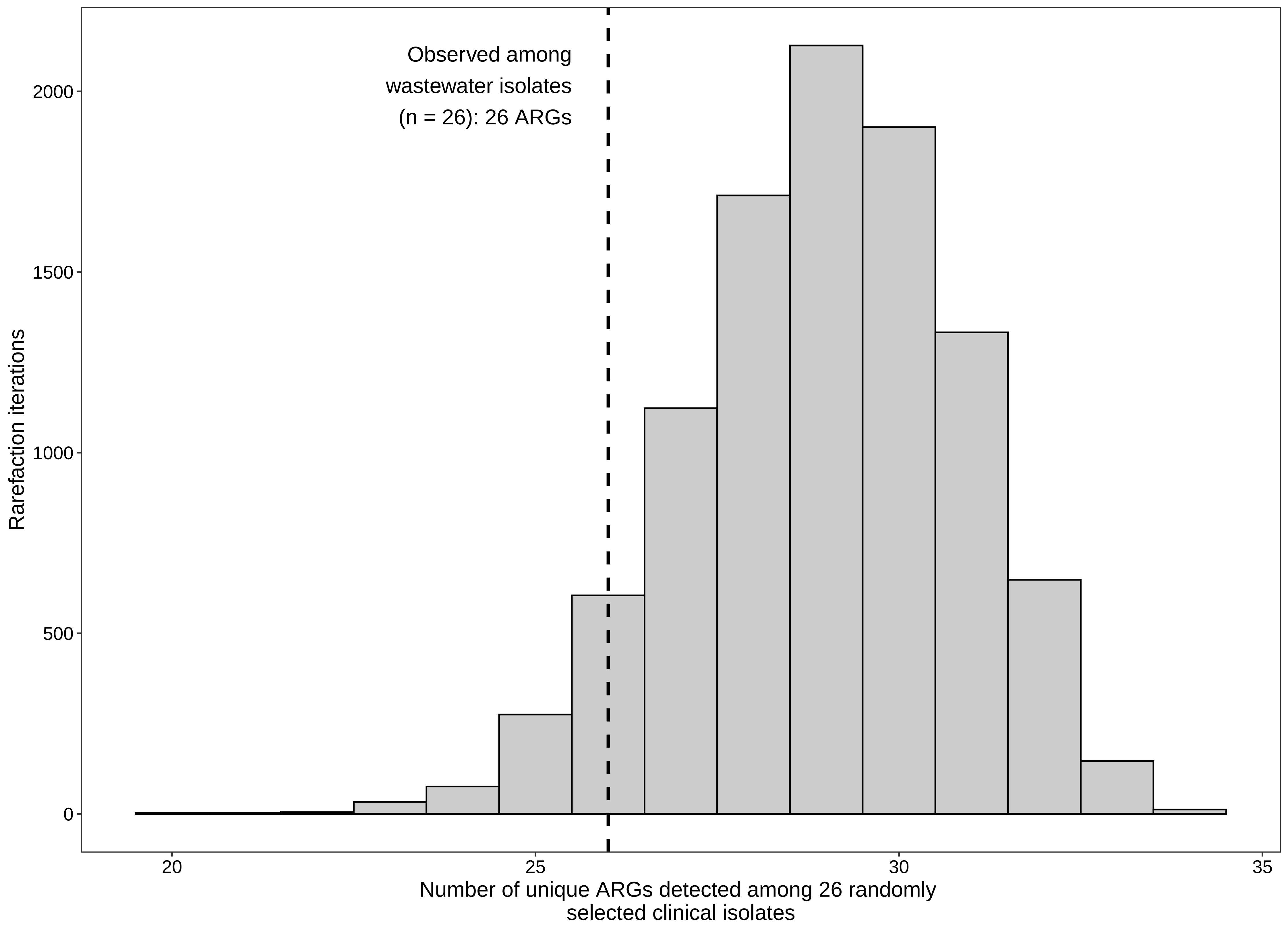
